# *Lactobacillus casei* Shirota intervention modulates the esophageal microbiome composition in Barrett’s esophagus

**DOI:** 10.1101/2025.05.12.25327438

**Authors:** Yonne Peters, Laura Ferrando, Chengliang Zhou, Rene te Morsche, Britt van der Leeden, Renske Cremers, Phuc Dat Le, Leander van Dijk, Ruud WM Schrauwen, Adriaan C Tan, Rachel S. van der Post, Peter van Baarlen, Peter D. Siersema, Annemarie Boleij

**Author notes:** Corresponding Author: Dr. Annemarie Boleij, Geert Grooteplein-Zuid 10, 6525 GA Nijmegen, The Netherlands. Shared first authorship (Yonne Peters, Laura Ferrando). shared last authorship (Peter D. Siersema, Annemarie Boleij).

## Abstract

**Objective:** Esophageal adenocarcinoma (EAC) and its precursor, Barrett’s esophagus (BE), are associated with a pro-inflammatory, Gram-negative-dominated esophageal microbiome. We investigated whether Lactobacillus casei Shirota (LcS) consumption could shift the microbiome towards a more Gram-positive profile in BE patients.

**Design:** In a single-arm intervention study, 23 BE patients used LcS twice daily for 4 weeks. Endoscopic biopsies from normal squamous epithelium (NSE) and metaplastic columnar epithelium (MCE), both before and after LcS-intervention were analyzed using 16S rRNA gene sequencing for microbiome composition, digital droplet PCR for Gram-positive to negative ratio, and fluorescence in situ hybridization to visualize Eubacteria and Firmicutes.

**Results:** LcS intervention increased Gram-positive Firmicutes in MCE *in situ* (p<0.01) while the overall abundance of Eubacteria was unaffected. Analysis at DNA level showed an increased Gram-positive to Gram-negative ratio post-LcS. The abundance of Gram-negative Proteobacteria lowered from 74% pre-LcS to 52% post-LcS, while the abundance of Gram-positive Firmicutes, including the genus *Lactobacillus*, increased from 20% pre-LcS to 31% post-LcS. Overall microbiota diversity significantly increased post-LcS (p=9.9778e-07). LcS intervention also resulted in an increased abundance in the BE associated taxa *Prevotella* and *Haemophilus* in both NSE as MCE.

**Conclusion:** LcS consumption significantly altered esophageal microbiome composition in BE patients, increasing Gram-positive taxa and overall diversity. These findings underscore the potential of probiotic-based therapeutic strategies aimed at EAC prevention. The concurrent rise in BE-associated taxa warrants careful consideration and further personalized studies to maximize benefits and minimize unintended consequences for optimal outcomes in BE patients.

**Significance of this study:** *What is already known on this subject?:*

- Epidemiological studies point to esophageal microbiota as a risk factor for BE.
- Gram-positive bacteria are closely associated with the normal distal esophagus, while Gram-negative bacteria (*Veillonella, Prevotella, Haemophilus, Neisseria*, and *Fusobacterium*) are increased in BE and EAC.
- LcS has been shown to have potent anti-tumor & anti-metastatic effects on colon and gastric cancer cells and to suppress chemically-induced carcinogenesis in animals.

*What are the new findings?:*

- The microbiota of both normal squamous epithelium and post-LcS samples show a higher microbiota diversity, indicating greater richness and evenness in the distal esophagus after LcS intervention.
- LcS intervention led, at the phylum level, to a significant increase in the relative abundance of health-associated, Gram-positive Firmicutes, increasing from 22% to 32% (\*\*\**p* ≤ 0.005). Gram-negative Proteobacteria showed a reduction from 72.0% to 54% (\*\*\**p* ≤ 0.005).
- Although LcS intervention led to an increase in Gram-positive Firmicutes, including *Lactobacillus*, it also resulted, in both normal squamous epithelium and metaplastic columnar epithelium, in an increased abundance of bacterial taxa associated with BE and EAC, including the genera *Prevotella* and *Haemophilus*.

*How might it impact on clinical practice in the foreseeable future?:*

- Our results suggest that the esophageal bacterial composition in BE patients can be modified by consumption of dairy products containing probiotics. Manipulation of microbiota aimed at altering microbiota composition to include higher abundances of health-associated Gram-positive taxa may provide a novel way for disease prevention and therapeutic intervention.

## Introduction

The incidence of esophageal adenocarcinoma (EAC) has been rising rapidly in Western countries over the past few decades(1). Because EAC is frequently detected at an advanced stage, patients with EAC have a dismal prognosis with a five-year survival rate of less than 20%(2). The only known precursor of EAC is Barrett’s esophagus (BE), with BE patients having a 30-125 fold increased risk of developing EAC(3). BE is characterized by the replacement of normal squamous epithelium (NSE) in the distal esophagus with metaplastic columnar epithelium (MCE). This metaplastic change is largely attributed to gastroesophageal reflux disease (GERD), affecting globally about 15-20% of the general population(4), resulting in injury of the squamous esophageal mucosa. However, not all patients with severe GERD develop BE(5).

While GERD is the main risk factor for BE, BE pathogenesis is complex and multifactorial. There is increasing evidence that the esophageal microbiome differs in patients with and without BE and may play a role in BE and EAC development(6). Once thought to be minimal, the esophageal microbiome is now recognized as having a diverse and stable bacterial population(7). In healthy individuals, the esophagus is predominantly colonized by carbohydrate-degrading, short-chain fatty-acid (SCFA)-producing Gram-positive bacteria, especially taxa of the genus *Streptococcus*(8–10). In contrast, BE is characterized by an increase in the relative abundance (RA) of disease-associated Gram-negative bacteria(3, 4)(taxa from the genera *Fusobacterium*, *Veillonella*, *Prevotella, Haemophilus, Neisseria,* and *Campylobacter*(11)) and a decrease in Gram-positive bacteria, relative to healthy individuals. This shift in BE suggests that a link might exist between the ratio of Gram-positive to Gram-negative taxa, and EAC development (12, 13). A hypothetical shift in Gram-positive to Gram-negative taxa ratio aligns well with the recently discovered competition model between two major bacterial groups or “guilds”, one guild of mainly Gram-positive bacterial taxa specialized in carbohydrate fermentation and production of the SCFA butyrate, and the other guild characterized by overrepresentation of mainly Gram-negative taxa associated with disease and antibiotic resistance, that distinguish human disease-from control cases(14). The relative increased abundance of disease-associated, often Gram-negative taxa observed in GERD may thus contribute to the inflammatory environment associated with BE and potentially influence its progression to EAC.

Probiotic administration has shown promise in modulating gut microbiome composition in gastrointestinal conditions. While research on probiotics in the context of BE is limited, studies in related conditions show interesting insights. For instance, probiotic consumption of Gram-positive taxa, such as members of carbohydrate-degrading and SCFA-producing genera *Lactobacillus*, *Bifidobacterium* and *Streptococcus*, may improve GERD symptoms(15). Probiotics may help restore bacterial balance by increasing the abundance of Gram-positive species, potentially counteracting the shift towards Gram-negative, frequently disease-associated taxa, and thus, fitting the abovementioned “two-competing-guilds” model correlating with health or disease. *In vitro* studies using a BE model have demonstrated that probiotic consumption can be associated with substantially lower expression of key biomarkers associated with progression of BE to EAC. Of particular interest is *Lactobacillus casei* strain Shirota (LcS), frequently used in probiotic dairy products(16), which has demonstrated potent anti-tumor, anti-metastatic, and anti-proliferative effects in various cancer models, including gastro-intestinal cancer(17–20). Although the specific impact of probiotics on the esophageal microbiome in BE patients requires further investigation, these preliminary findings indicate potential for manipulation of esophageal microbiota composition to improve BE management and EAC prevention.

We hypothesized that consumption of probiotic LcS by BE patients might result in an increase of Gram-positive taxa and a decrease of Gram-negative disease-associated taxa in the esophageal microbiome. To test this hypothesis, we invited BE patients undergoing surveillance endoscopies to consume a probiotic drink containing LcS twice daily for 4 weeks. The goal of our study was to measure and monitor changes in bacterial composition following LcS intervention in MCE and NSE of BE patients, to unravel potential implications in BE pathogenesis and propose novel EAC prevention strategies.

## Methods

### Study design

We performed a single-arm, interventional pilot study to evaluate the ability of changing the esophageal microbiota by ingestion of a fermented milk drink containing LcS in patients with BE with confirmed MCE with intestinal metaplasia. We analyzed the microbial composition of biopsies obtained from NSE and BE regions (MCE) before and after use of LcS to identify potential alterations in mucosal microbiota composition. In addition, the ability of LcS to colonize the esophagus and its impact on the Gram-positive to Gram-negative ratio was investigated by DNA sequence profiling of bacteria and fluorescent *in situ* microscopy analysis of biopsy sections. The study was in accordance with the Declaration of Helsinki, the code of conduct for Health Research, and was approved by ethics committee CMO Arnhem Nijmegen (NL59072.091.16) and registered in the Netherlands Trial Register (NL-OMON43164). All the participants provided informed written consent.

### Participants

Study participants who had been referred for a surveillance upper endoscopy for BE were recruited from two hospitals (Bernhoven Hospital, Uden, the Netherlands and Radboud university medical center, Nijmegen, the Netherlands). Individuals aged 18 years or older with a Barrett segment of at least 2 cm with histologically proven columnar-lined epithelium (metaplasia) without dysplasia, were eligible to participate. Exclusion criteria included probiotic or antibiotic use within the last 3 months before baseline, infection of the oral cavity, vegetarian or gluten-free diet, lactose intolerance, immunodeficiency disorders, bleeding disorders, *Helicobacter pylori* infection, previous gastric or esophageal surgery, and other coexistent esophageal diseases (e.g. varices or reflux esophagitis).

### Intervention

Patients received a fermented probiotic drink (Yakult: 65ml; Yakult Nederland BV, Amstelveen, the Netherlands) containing a minimum of 6.5·10^9^ colony-forming units (CFU) LcS/bottle twice-daily for four weeks. Other Yakult ingredients were water, skimmed milk powder, glucose/fructose syrup, sugar, maltodextrin, and flavorings. Fourteen fresh bottles of Yakult were delivered by a courier service to the participants’ home addresses every week. Patients were asked to be fasting 15 minutes before and after drinking the bottle. Compliance was assessed by a patient diary and registration of the remaining unopened bottles by the courier service. During the intervention period, patients were not allowed to take other probiotics or antibiotics. Patients receiving antibiotic therapy were excluded from the study.

### Sampling and analysis procedures

The primary outcomes were (i) the effect of LcS on the Gram-positive to Gram-negative ratio in the esophagus, and (ii) the effect on the diversity and composition of the microbiota in NSE and MCE epithelium. Secondary outcomes were the impact of LcS on endoscopic and histologic inflammation. The additional methods concerning sampling procedures, tissue processing and scoring, quantitative PCRs, microbiota analysis and statistical analysis can be found in **Supplemental methods** in **Supplemental files**.

## Results

### Patient enrollment and baseline characteristics

A total of 31 patients were enrolled and underwent baseline endoscopy. Of these, 25 patients who met inclusion criteria and had the necessary number of study biopsies taken started the 4-week Yakult intervention. Twenty-three patients completed the study, including the second endoscopy (**Figure 1**). Baseline characteristics are shown in **Table 1**. Analysis performed on patient samples are listed in **Supplemental table S1.** Most patients were male (73.9%), with a median age of 68 years (IQR 59–71). Median BE segment length was Circumferential (C) 2 cm and Maximum (M) 4 cm (IQR C0–5, M3–7). All patients were on acid suppression therapy with either proton pump inhibitors (PPIs) or H2 receptor antagonists. No patients reported reflux symptoms at baseline and endoscopic evaluation showed no signs of inflammation in any of the patients. Histopathological evaluation of clinical FFPE biopsies confirmed presence of MCE with intestinal metaplasia in the BE segment without dysplastic changes in all patients at baseline.

**Figure 1.**
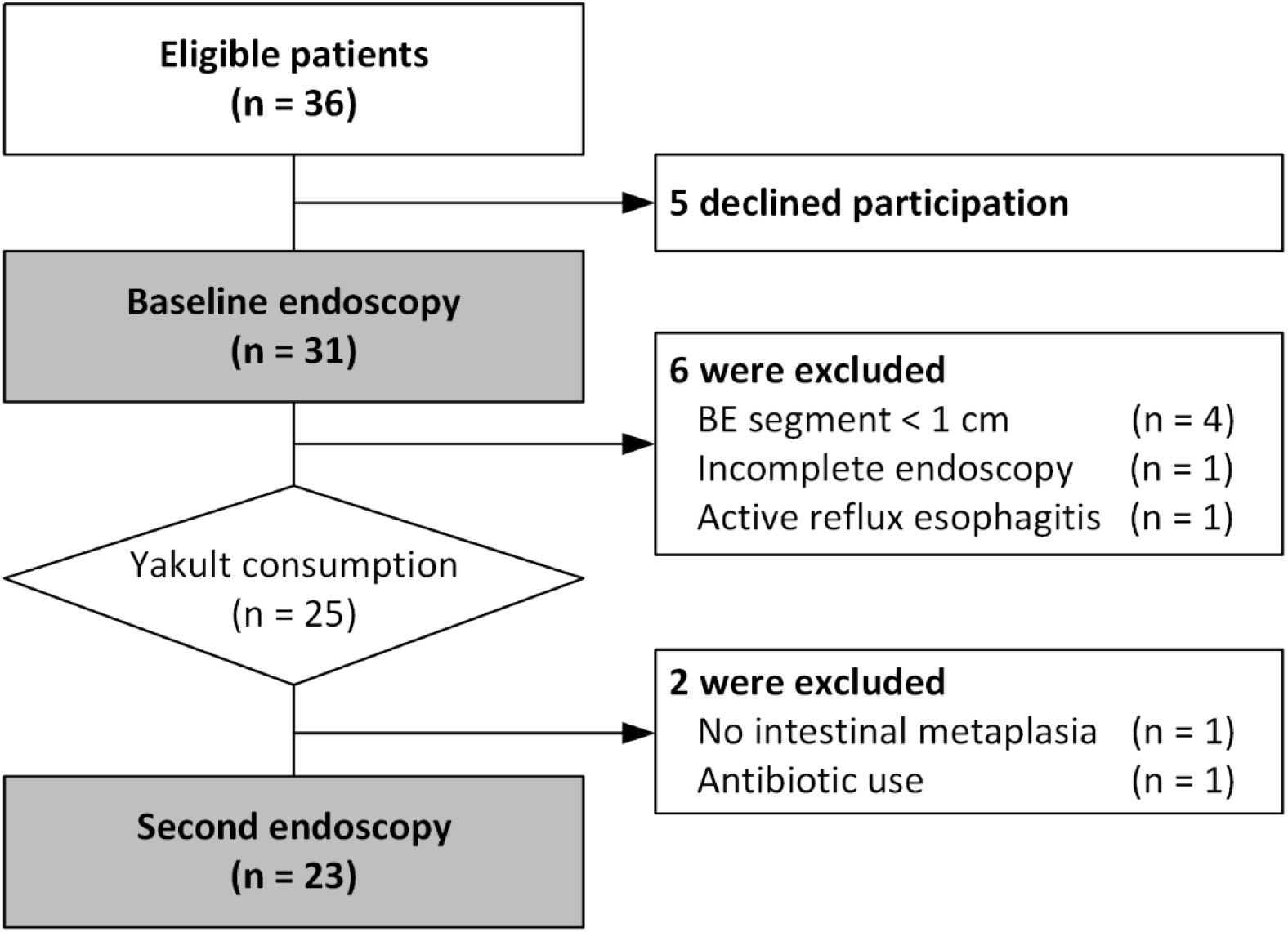
Study population

**Table 1.**
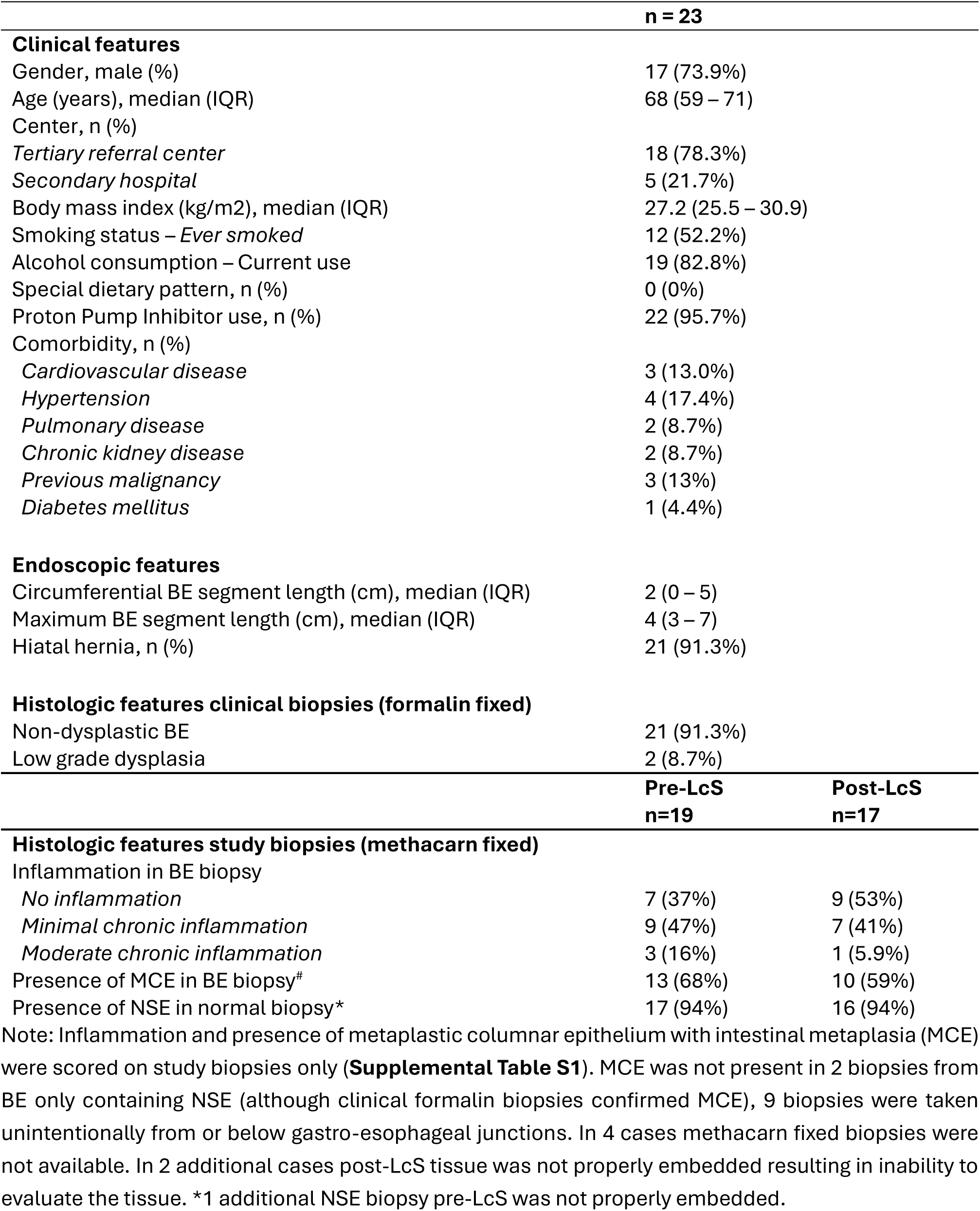
Patient characteristics.

### Clinical evaluation

Self-reported adherence to Yakult regimen was high, with 96% of patients reporting >90% compliance (1 diary missing). No serious adverse events were reported during the study period. Most common minor side effects were diarrhea (n=2) and mild bloating (n=1). Following the 4-week Yakult intervention, the second endoscopy revealed no significant changes in endoscopic features or clinical characteristics compared to baseline. The absence of initial reflux symptoms or endoscopic inflammation made it not possible to evaluate any potential improvements due to Yakult consumption.

### Histopathological evaluation and *In situ* detection of bacteria

Histopathological evaluation of the endoscopically guided Methacarn fixed study biopsies, intended for microbial *in situ* bacterial evaluation, were scored for presence of MCE and inflammation. In 68% and 59% of BE segment study biopsies MCE with intestinal metaplasia was detected pre- and post-LcS intervention, respectively. Minimal or moderate chronic inflammation was observed in 63% and 47% of MCE biopsies pre- and post-LcS intervention, respectively (p>0.05; **Table 1**). Histologic inflammation was not observed in any of the NSE biopsies. We evaluated the *in situ* presence of Eubacteria, Gram-positive Firmicutes, and Gram-negative Bacteroidetes and Gammaproteobacteria using fluorescence microscopy. Representative images of histology, Fluorescent *in situ* hybridization (FISH) for Eubacteria and Firmicutes are shown in **Figure 2** for MCE, with corresponding controls and NSE biopsies in **Supplemental Figure S1 and S2**. No differences were observed in the number of Eubacteria signals per mm apical region in both MCE (median 2.19, and 2.93), and NSE biospies (median 1.35 and 0.96) pre- and post-LcS, respectively. A significant increase in Firmicutes signals post-LcS was observed per mm apical region for MCE biopsies (median 0.32 vs 1.67; ** p < 0,01 Wilcoxon Signed Rank test). A similar trend was observed in NSE biopsies, but this was not significant (median 0.4 vs 0.86) (**Supplemental Figure S3**). The Gram-negative bacterial taxa Bacteroidetes and Gammaproteobacteria were detected in a minimal number of samples at low abundance pre- and post-LcS.

**Figure 2.**
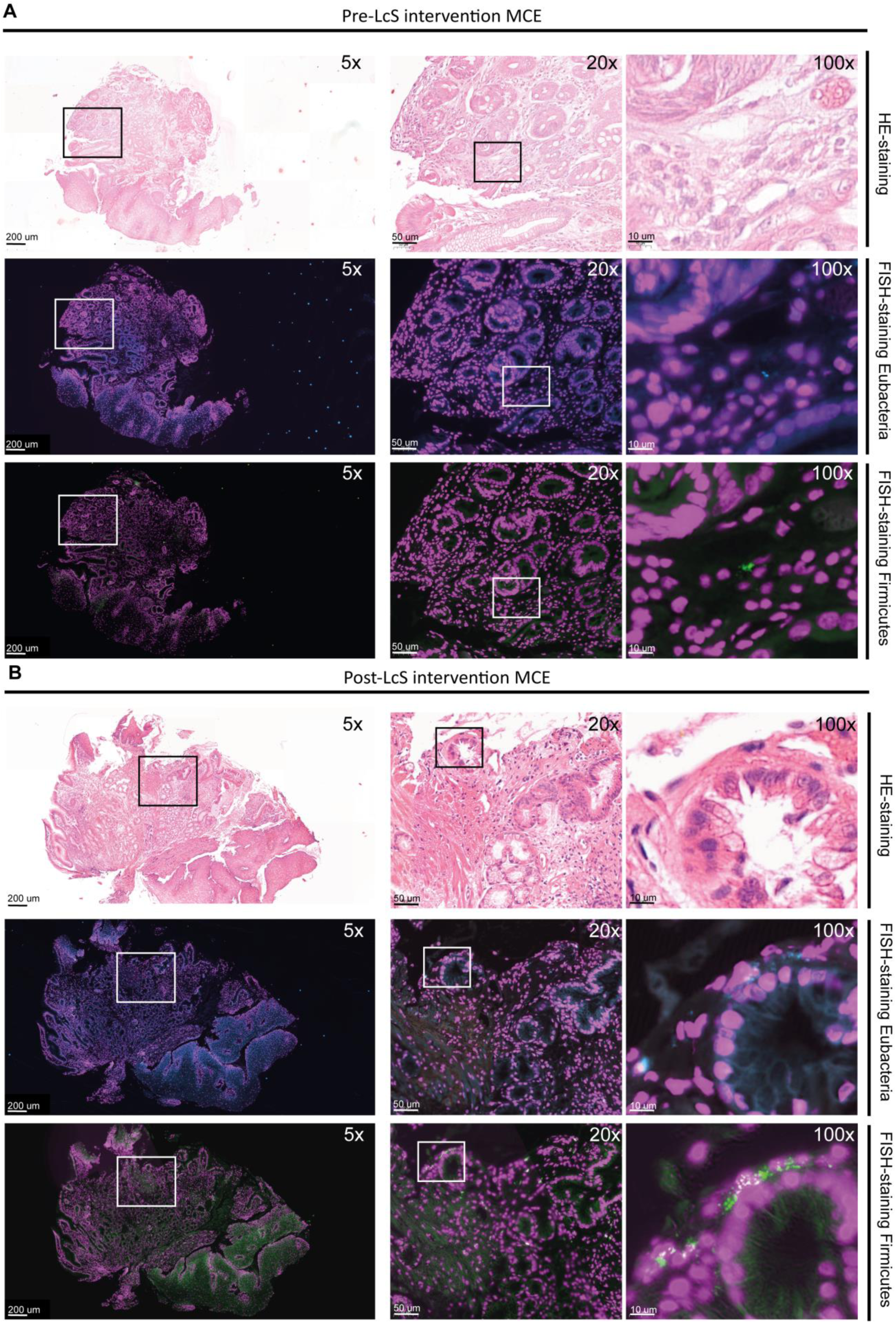
FISH detection of bacteria in MCE biopsies pre- and post-LcS intervention. Representative H&E staining and corresponding FISH images of Eubacteria (cyan) and Firmicutes signals (green) detected in MCE biopsies of 1 case with high Eubacteria and Firmicutes presence pre-LcS intervention (A) and post LcS intervention (B) at 5x, 20x en 100x zoom. Nuclei are visible in magenta (DAPI stain).

### Gram-positive to Gram-negative ratio in MCE and NSE biopsies pre- and post LcS intervention

Our working hypothesis was that the consumption of Gram-positive LcS probiotics would lead to an increase in the RA of Gram-positive taxa and a reduction in the RA of Gram-negative taxa. The Gram-positive to Gram-negative ratio measured with droplet digital PCR (ddPCR), a quantitative PCR method that counts DNA target copies in nanoliter-sized droplets, showed a higher RA of Gram-negative bacteria in both NSE and MCE regions pre-LcS (**Supplemental Figure S4A**). Post-LcS intervention, the Gram-positive to Gram-negative ratio increased from 0.42 to 1.02 (\*\*\**p* ≤ 0.005) in NSE regions and 0.48 to 1.11 (P>0.05) in MCE regions, covering 90% and 75% of the pairs respectively (**Supplemental Figure S4B**). We were unable to confirm specific colonization of the esophagus by the LcS strain in neither NSE nor MCE biopsies after twice daily consumption.

### General characteristics of esophageal microbiota in BE-patients at baseline

To assess whether the intake of LcS could alter esophageal microbiota of BE patients, we compared the microbial composition, alpha diversity and RA between MCE and NSE biopsies. At the phylum level, BE patients showed a high abundance of Gram-negative Proteobacteria (55.1%), followed by Gram-positive Firmicutes (23.5%), Actinobacteria (4.6%) and Bacteroidetes (3.5%). The remaining taxa included Fusobacteria, Epsilonproteobacteria, and Patescibacteria. Among the Proteobacteria, the most common genus was *Mesorhizobium* (32.1%), followed by *Xanthobacteraceae* (12.2%%) (specifically *Bradyrhizobium*) and *Sphingomonas* (3.5%) (**Supplemental Figure S5A**). Other common taxa included members of the Gram-positive Firmicutes such as *Streptococcus* (16.6%) and *Rothia* (Actinomycetota) (3.0%) and less abundant taxa such as the Gram-positive genus *Staphylococcus* (2.7%), and the Gram-negative genera *Prevotella* and *Reyranella* (both around 2%) (**Supplemental Figure S5B**). The Shannon index, a standard measure of diversity that considers both the number of taxa and their proportional abundance, was significantly higher in NSE compared to MCE at the genus level (*p* = 2.0e-06) (**Figure 3A**). To test the hypothesis that BE is associated with a higher RA of Gram-negative taxa, we compared the RA of the Gram-negative phylum Proteobacteria and the Gram-positive phylum Firmicutes between NSE and MCE biopsies. At the phylum level, no significant differences were observed between RA of Proteobacteria (NSE= 66.3% vs MCE= 62.8%) and Firmicutes (NSE=26.0% vs RA MCE=25.5%) in MCE vs NSE biopsies. In addition, we performed a partial-RDA analysis for tissue origin (MCE vs NSE) and corrected this for the covariates LcS-intervention and patients because these could at least partially explain variation in microbiota RA. The tissue origin MCE or NSE, contributed 25.5% to the total variation in bacterial RA (*p* = 0.002), showing that the bacterial composition differed significantly between NSE and MCE of the esophagus (**Figure 3B**). The genera at highest differential RA in NSE included *Staphylococcus*, *Chitinophaga*, *Reyranella*, *Hephaestia*, *Pseudolabrys*, (RDA values 0.70-0.94). In contrast, the genera *Rothia*, *Acinetobacter, Actinomyces, and Granulicatella*, were at higher RA in MCE (RDA values −0.48 to −0.38; **Figure 3C** and **3D**).

**Figure 3.**
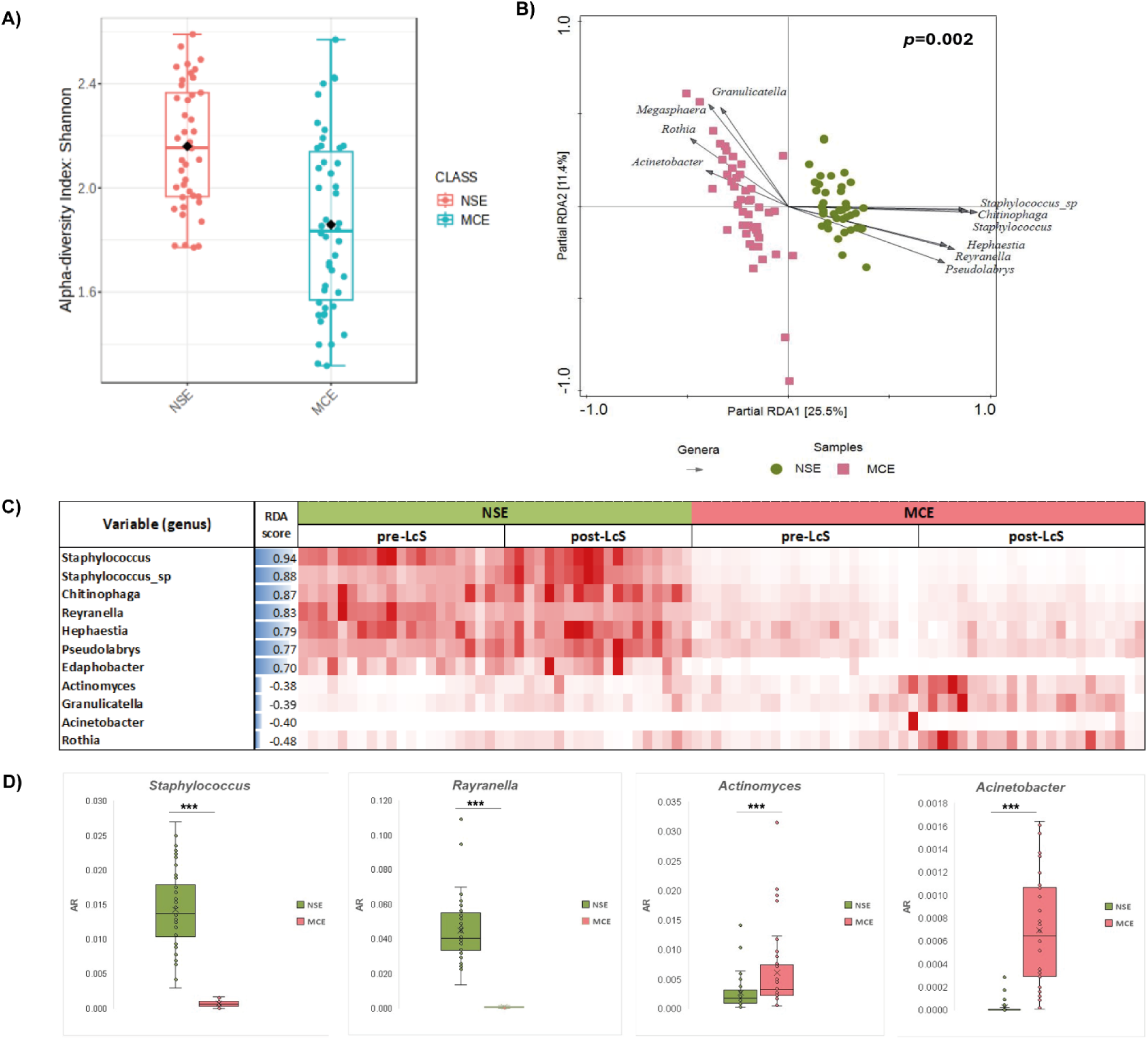
RDA and RA reveal the most discriminative genera in MCE vs NSE. **A)** Differences in the Shannon α-diversity index of microbial communities at genus level between esophageal locations (NSE and MCE) **B**) Partial Redundancy analysis (RDA) of genera adjusted for patients and LcS-intervention of oesophagal region samples; the x-axis separates NSE from MCE and explains 25.5 % of the microbial composition differences observed; microbial community composition sampled from the two regions in the oesophagus was significantly different (*p* = 0.002). The top 10 most discriminative genera are indicated with black arrows. The direction of arrows correlates with location, and arrow lengths with correlation strength. **C**) The relative abundance (RA) of the most discriminative genera displayed as heatmap. **D**) Boxplots show RA of the most discriminative genera. \*\**p* ≤ 0.01, *** *p* ≤ 0.005 Multiple Linear Regression adjusted with Covariates: treatment and patients.

### Higher bacterial diversity in BE and squamous epithelium post-LcS intervention

Next, we evaluated specific changes in microbial diversity comparing pre- and post-LcS biopsies from NSE and MCE regions. Administration of LcS increased bacterial richness in both NSE and MCE esophageal locations, with significant differences observed at the genus level (*p*= 2.8e-05) (**Figure 4A**, **4B**, **Supplemental Table S3**). Multivariate PERMANOVA based on Bray–Curtis distances revealed distinct microbial communities at the genus level that occurred at significantly differential RAs between NSE and MCE (*p* = 0.001) (**Figure 4C**), and between pre- and post-LcS (**Supplemental Table S4**). Principal component analysis (PCA) corroborated these findings, showing that nearly 60% of the variation was captured by the first and second principal component (PC1&2) (**Supplemental Figure S6**). Both the two esophageal regions (NSE vs MCE) and the intervention (pre- vs post-LcS) contributed statistically significant to the separation of microbiota (MCE vs NSE, *p*= 0.001; pre-LcS vs post-LcS, *p*= 0.001, **Supplemental Table S3**). The genera that contributed most to the differences in microbiota composition between NSE and MCE regions included *Prevotella*, *Megasphaera*, and *Actinomyces*, the principal discriminating genera in MCE. In contrast, bacteria belonging to the genera *Staphylococcus*, *Reyranella*, and *Chitinophaga* were more dominant in NSE (**Supplemental Figure S6**).

**Figure 4.**
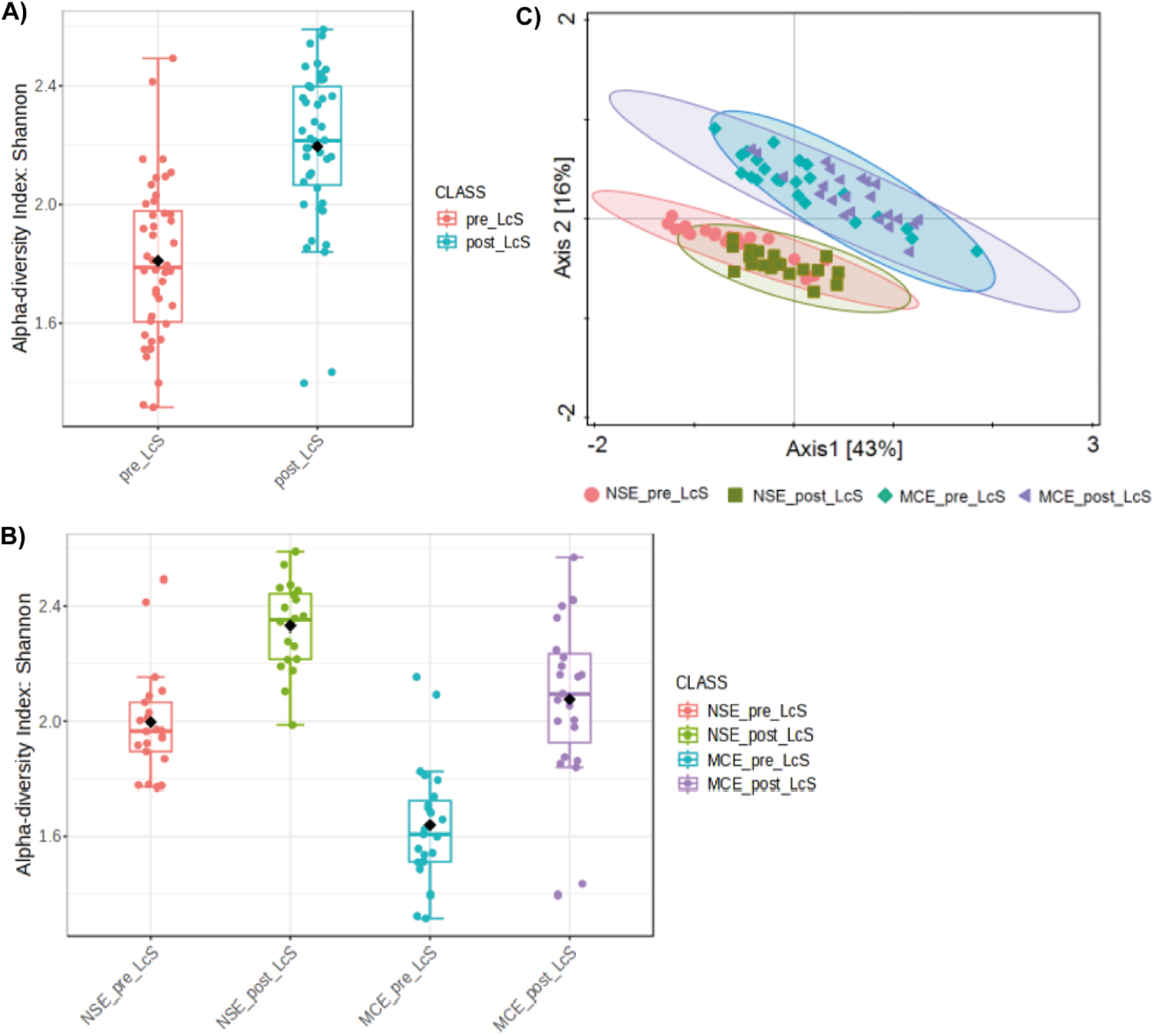
Differences in α-diversity and b-diversity of MCE vs NSE pre- and post-LcS intervention. Differences in the Shannon α-diversity index of microbial communities at genus level: **A**) before and after administration of LcS (pre-LcS and post-LcS) **B**) pre-LcS and post-LcS for each esophageal region. **C**) Principal coordinate analysis (PCoA) visualizing differences in microbiota composition at genus level associated with sampled esophageal regions and pre- or post-LcS intervention sampling time points. PcoA was generated using Bray–Curtis distance at genus composition level. The experimental groups were distinct by esophageal tissue origin (NSE vs MCE) and for intervention (pre-vs post-LcS).

### LcS intervention increased specific Gram-positive taxa and reduced specific Gram-negative taxa

The administration of LcS led to a significant increase in the RA of Gram-positive Firmicutes at the phylum level, showing an increase from 22% to 32% in both NSE and MCE biopsies (Tukey’s HSD: \*\*\**p* ≤ 0.005, p<0.05, respectively). Gram-negative Proteobacteria showed a reduction from 72.0% to 54% (Tukey’s HSD:\*\**p* ≤ 0.01 for both NSE and MCE) (**Figure 5A**). Among Firmicutes, the RA of the genus *Lactobacillus* had increased from 0.01% to 0.03% (Tukey’s HSD: \*\*\**p* ≤ 0.005 for MCE only) (**Figure 5B**). The significant increase in Firmicutes, and reduction in Proteobacteria was only observed in patients included at the Radboudumc hospital and not at the Bernhoven s**i**te **(Supplemental Figure S7)**. These findings corroborate the findings from the Gram-positive to Gram-negative ratio assessed with ddPCR and the *in situ* increased detection of Firmicutes post-LcS.

**Figure 5.**
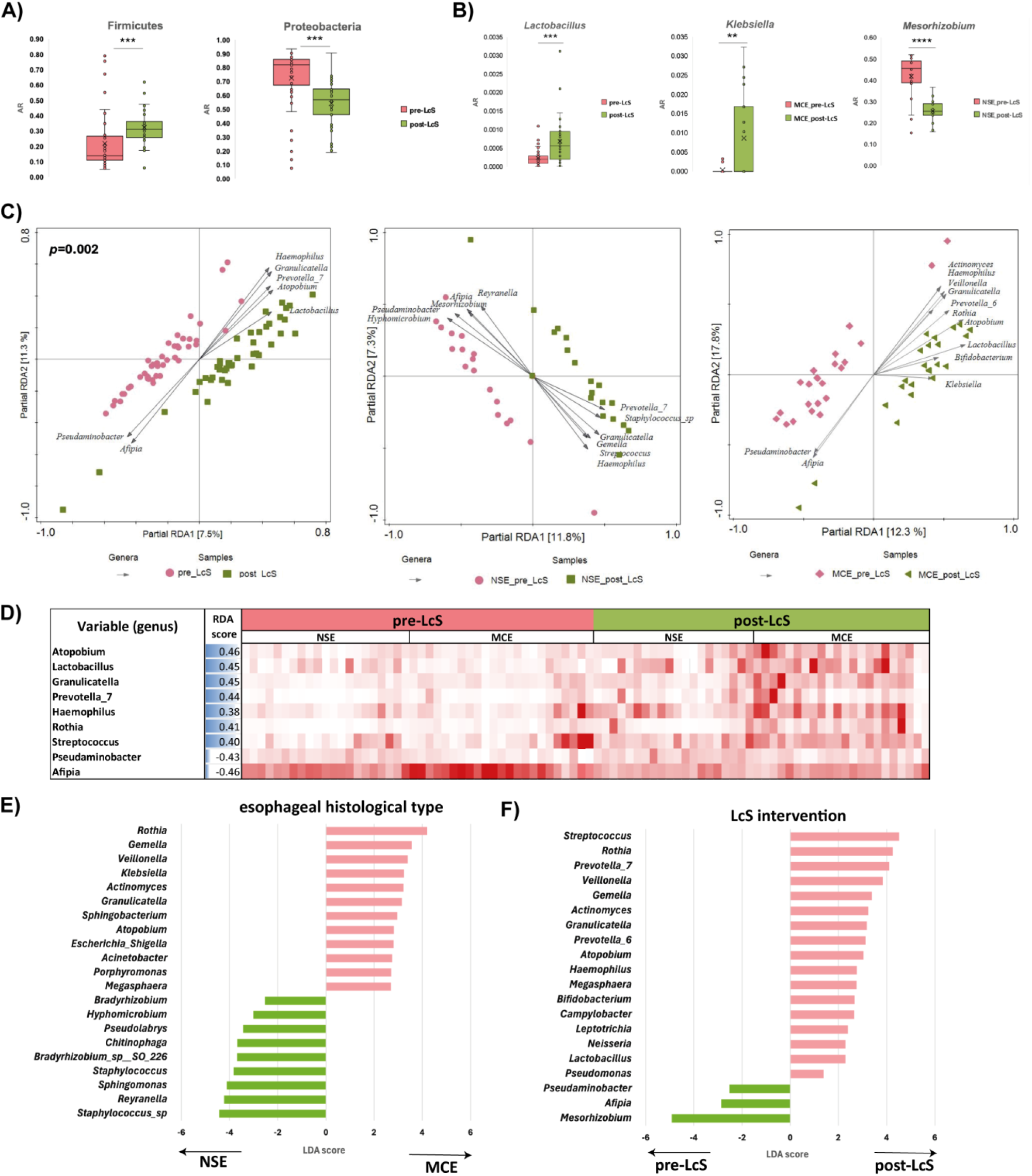
RDA analysis and RA identify the most discriminative genera pre- and post LcS intervention. **A**) The relative abundance (RA) of the phyla Firmicutes and Proteobacteria before and after LcS intervention. **B)** RA of the most discriminative genera *Lactobacillus*, *Mesorhizobium* in NSE and *Klebsiella* in MCE biopsies pre-versus post-LcS intervention **C**) Partial-RDA of genera grouped for NSE and MCE toghether comparing pre-with post-LcStissue of origin (MCE vs NSE) and intervention (pre-vs post-LcS) separately. The RDA1 axis separates pre-LcS from post-LcS and explains 6.5 % of observed differences in bacterial composition. Bacterial community composition changed significantly due to intervention with LcS (*p* = 0.002). The top 10 most distinctive genera are represented with black arrows. Arrow directions correlate with esophageal location, arrow lengths with correlation strength. **D**) RA of the most discriminative genera displayed as heatmap. (**E**) Rank correlation analysis using Linear Discriminant Analysis (LDA) Effect Size (LEfSe) was performed to compare microbial features, candidate biomarkers, at genus level between esophageal regions (NSE and MCE) and (**F**) before and after intervention with LcS. Phylum with LEfSe scores >2.5 or <−2.5 and an FDR-corrected *p* < 0.05 were considered significant. \*\**p* ≤ 0.01, *** *p* ≤ 0.005 Multiple Linear Regression adjusted with Covariates: esophageal regions and patients.

The impact of LcS intervention on microbiota composition at the genus level was assessed using partial-RDA, adjusted for patients and tissue origin (MCE vs NSE phenotype; detailed Methods in supplement) (**Figures 5C** and **D**). Partial-RDA revealed that the intervention (LcS consumption) explained 7.5% variation in microbiota composition of the esophageal microbiota pre- and post-LcS intervention. Post-LcS intervention, genera that contributed strongest to differences in microbiota composition in the esophageal tissue origin included *Lactobacillus*, *Rothia*, *Streptococcus*, *Veillonella*, and *Klebsiella* (**Figures 5C** and **D**).

The genera contributing most to the differential microbiota composition across the NSE and MCE regions was investigated. In the NSE region, LcS consumption led to a significant reduction in Gram-negative *Mesorhizobium*, from 41% to 25% (\*\*\*\**p* ≤ 0.001) (**Figure 5B**). Although LcS intervention had led to a decrease in Gram-negative Proteobacteria, it also resulted in an increase in the taxa *Prevotella* and *Haemophilus*, in both the NSE and MCE regions. Members of the genus *Klebsiella* also showed a significant increase in MCE biopsies(**p<0.01), but remained at low RA (**Figure 5B, Supplemental Figure S7**). Next, we employed Linear discriminant analysis Effect Size (LEfSe), specifically designed for biomarker discovery rather than multivariate analysis of RA, across sampling regions. LEfSe determines the taxa most likely to explain differences in microbiota composition between NSE and MCE by coupling standard tests for statistical significance with additional Linear Discriminant Analysis (LDA) tests, to improve biological consistency and impose a given effect size(21). LEfSe was used to identify taxa associated with BE by comparing microbiota composition between NSE and MCE biopsies, as well as to identify (biomarker) taxa associated with impact of LcS consumption. LefSe analysis corroborated our previous findings, uncovering Firmicutes as a biomarker post-LcS intervention (LDA score= 4.66, *p* ≤ 0.005, **Table S4**). Members of the phyla Proteobacteria (Pseudomonadota) and Fusobacteria (Fusobacteriota) were listed as enriched candidate biomarkers pre-LcS intervention (**Supplemental Figure S6, Table S4**). According to LEfSe analysis, members of the genera *Staphylococcus*, *Reyranella*, and *Chitinophaga* were biomarkers associated with NSE regions, while members of the genera *Streptococcus, Rothia* and *Prevotella_7* were designated as post-LcS intervention biomarkers (**Figure 5E** and **F**).

## Discussion

Our study highlights significant differences in bacterial composition of NSE and MCE in BE patients, accounting for 25.5% of the observed variation in bacterial composition. LcS use led to notable shifts in the esophageal microbiota composition, including increased microbial diversity in both epithelial regions and a significant increase in the Gram-positive/Gram-negative ratio. Specifically, we observed a marked increase in the RA of Gram-positive Firmicutes (from 22% to 32%) alongside a reduction in Gram-negative Proteobacteria (from 72% to 54%). While *Lactobacillus* abundance increased significantly post-LcS intervention, taxa previously associated with BE and EAC, such as *Prevotella* (*22*), also showed increased abundance in both epithelial regions. These findings suggest that although LcS modulates the esophageal microbiota toward a more diverse and potentially beneficial composition, undesired increases in RA of disease-associated taxa warrant further investigation into clinical significance.

Our finding that within the same individual, esophageal squamous and metaplastic epithelium exhibit substantially different bacterial compositions underscores the importance of the local epithelial environment in shaping the esophageal microbiome. Higher bacterial diversity, especially of Gram-positive taxa, were features of healthy esophageal squamous epithelium, whereas Gram-negative bacteria appeared to be abundant in metaplastic epithelium. This observed shift from Gram-positive to Gram-negative predominance in BE is of clinical relevance. The outer membrane of Gram-negative bacteria possess lipopolysaccharide (LPS), potent inducers of inflammation, that may contribute to the metaplasia-dysplasia-carcinoma sequence observed in BE(12). Furthermore, certain Gram-negative species found in BE, such as *Campylobacter concisus*, produce toxins that damage epithelial cells and potentially contribute to carcinogenesis (23) (24). More generally, progression from healthy normal squamous epithelium in the esophagus to BE-associated metaplastic columnar epithelium and subsequently to EAC is characterized by an increased RA of Gram-negative bacterial species (12) including disease-associated species from the genera *Campylobacter* and *Fusobacterium* (25) (26) (27). Conversely, Gram-positive bacteria, particularly certain *Streptococcus* and *Lactobacillus* species, of which we assayed the strain *L. casei* Shirota, may play protective roles in esophagus through production of bacteriocins, short-chain fatty acids (SCFAs) from the breakdown of complex carbohydrates, and competition with pathogenic species (7). The SCFAs lactate and acetate modulate immune response and host tolerance, and contribute to competitive capacity of Gram-positive bacteria against pathogenic, often Gram-negative, microorganisms (28) (29). These contrasting ecological properties of Gram-negative and Gram-positive taxa align with the recently discovered model of two competing guilds, one mainly containing Gram-negative taxa that include common disease-associated species and lineages, and another one containing Gram-positive taxa associated with anaerobic fermentation, butyrate production and health (14). Interestingly, we found that Gram-negative *Mesorhizobium*, overrepresented in patients with GERD (30, 31) and at high RA in squamous epithelium of our patients, significantly decreased after LcS consumption.

In the metaplastic esophageal region, we observed increase RA of typical intestinal taxa like *Klebsiella* and *Prevotella*, possibly driven by gastro-esophageal reflux. The high glucose content in probiotic Yakult drink, may stimulate growth of pro-inflammatory bacteria and decrease capacity of mucosal tissues to regulate epithelial integrity and immunity (32). Increased RA of *Prevotella*, a taxon associated with dysbiosis and cancer (27), warrants further attention, as previous studies have linked *Prevotella* to inflammatory conditions and tissue damage through the production of pro-inflammatory mediators(33, 34). Homolactic fermentation of glucose by LcS and other Gram-positive bacteria, such as *Streptococcus*, potentially have a dual effect: modulating host immunity via lactate and acetate, and decreasing pH(35, 36). While RA of the genus *Lactobacillus* increased significantly following LcS intervention, its overall RA remained low (0.01% to 0.03%); increased RAs of Gram-positive *Rothia* and *Streptococcus* contributed strongly to the significant increase in Firmicutes. In this study, we were unable to confirm colonization with LcS using an LcS specific assay, suggesting that its direct colonization may not be the primary mechanism of action. To optimize clinical treatment, supplementation of LcS in drinks with low glucose content together with acid-inhibiting agents like proton pump inhibitors (PPIs), might enhance probiotic benefits including reducing RAs of disease-associated Gram-negative bacteria.

The findings of our study highlight clinical implications regarding the role of the esophageal microbiome in BE. First, microbial composition within metaplastic Barrett’s epithelium may serve as biomarker to identify patients at higher risk of progression to dysplasia or EAC. Specifically, increased abundance of taxa such as Proteobacteria and *Prevotella* was associated with inflammation and dysbiosis, possibly indicative of disease progression. Higher abundance of *Leptotrichia* (phylum Fusobacteria) was proposed by others as candidate biomarker of neoplastic esophageal mucosa(22). Changes in microbiota composition contributing to, or resulting from carcinogenic processes have positioned microbiome composition, especially RAs of Gram-positive- and Gram-negative taxa, as biomarker for disease progression and therapeutic targets (37).

The observed effects of LcS intervention on microbiome diversity and Gram-positive/Gram-negative ratio suggest potential of manipulating microbiota composition in managing BE. By enhancing microbial diversity and improving balance between Gram-positives and Gram-negatives, LcS-based therapies could mitigate inflammation and support epithelial integrity, reducing EAC risk. In a BE organoid model, L-lactate production by *Streptococcus* and *Lactobacillus* reduced EAC proliferation and modulated immune and inflammatory responses(36). Therapeutic modulation of the esophageal microbiota, through probiotics like LcS or other targeted interventions, thus represents a promising adjunctive strategy for BE management, complementing current approaches aimed at halting or reversing disease progression. These findings align with emerging evidence linking carefully designed microbiome-targeted interventions (for example, probiotic formulations low in glucose) to improved gastrointestinal health and cancer prevention(38).

This study included a thorough microbiome analysis using 16S rRNA sequencing, ddPCR, and FISH, providing a comprehensive view of the esophageal microbiota in BE patients. Studying squamous and metaplastic epithelium within the same subjects offered valuable insights into personalized, region-specific microbial differences. Furthermore, the within-subject design allowed for robust assessment of changes induced by LcS intervention. Still, limitations should be considered when interpreting the results. The relatively small sample size may limit generalizability of the findings. We noted differences in microbiome composition between patients included in different hospitals, underscoring that a more diverse study population is necessary to generalize conclusions. The intervention duration of four weeks, while sufficient to observe some changes, may not fully capture long-term effects of probiotic consumption on the esophageal microbiome. Additionally, the lack of a control group without probiotic consumption makes it challenging to distinguish between LcS intervention effects and potential confounding factors or natural fluctuations over time.

Future research directions could address these limitations and expand our understanding of the esophageal microbiome in BE. Studies with larger sample sizes will provide greater statistical power to detect subtle microbial composition shifts. Longer intervention periods could incorporate multiple clinical outcome measurements, possibly including follow-up to assess BE progression to EAC, to help determine whether microbiome composition predicts EAC development in BE patients. If such predictive patterns are identified, research could focus on whether modulating the microbiome through targeted interventions could slow or even, prevent progression to EAC. Finally, mechanistic studies to elucidate the underlying processes driving microbial shifts in BE and their potential role in disease progression, including impact of specific bacterial (toxic) metabolites and host immune responses, are needed.

In conclusion, LcS consumption led to an increase in Gram-positive taxa and microbial diversity together with a decrease in Gram-negative taxa in BE. The concurrent rise in specific disease-associated taxa that thrive on simple sugars such as *Prevotella*, *Haemophilus* and *Campylobacter* suggested that formulation of probiotic drinks low in simple sugars is desired. Our findings underscore the potential of microbiome modulation, by stimulation of health-associated Gram-positive SCFA-producing taxa such as *Lactobacillus* and *Streptococcus*, as a possible therapeutic strategy in BE management and EAC prevention.

## Competing interests and funding

PDS received unrestricted grants from Pentax, Fuji Film, Norgine, MicroTech, Magentiq Eye, AstraZeneca, Sanofi, and in the advisory board of Sanofi (The Netherlands). The study was partly funded by Yakult Europe B.V. for an amount of 40.000 euro. Yakult Europe B.V. did not have any input in the design, execution or interpretation of the outcome of the study.

## Patient & public involvement

Patients or the public were not involved in the design of the study as the study protocol was designed in 2017 and patient or public involvement was at that time not needed.

## Abbreviations used in this paper

LcS: Lactobacillus case Shirota
preLcS: before administration of LcS
postLcS: after administration of LcS
BE: Barrett’s esophagus;
EAC: esophageal adenocarcinoma
NSE: normal squamous epithelium
MCE: metaplastic columnar epithelium
RA: relative abundance
CI: confidence interval
HGD: high-grade dysplasia
IQR: interquartile range
SD: standard deviations

## Data Availability

All data produced in the present study are available upon reasonable request to the authors and microbiome data are available online at ENA under accession number PRJEB89339 and will become available when accepted for publication.

https://www.ebi.ac.uk/ena/browser/view/PRJEB89339

## Notes

### Clinical Trial

NL-OMON43164

### Author Declarations

The study was in accordance with the Declaration of Helsinki, the code of conduct for Health Research, and was approved by ethics committee CMO Arnhem Nijmegen (NL59072.091.16) and registered in the Netherlands Trial Register (NL-OMON43164). All the participants provided informed written consent.

## References

1. Arnold M, Laversanne M, Brown LM, Devesa SS, Bray F. Predicting the Future Burden of Esophageal Cancer by Histological Subtype: International Trends in Incidence up to 2030. The American journal of gastroenterology. 2017;112(8):1247–55.

2. Pohl H, Welch HG. The role of overdiagnosis and reclassification in the marked increase of esophageal adenocarcinoma incidence. Journal of the National Cancer Institute. 2005;97(2):142–6.

3. Peters Y, Al-Kaabi A, Shaheen NJ, Chak A, Blum A, Souza RF, et al. Barrett oesophagus. Nature Reviews Disease Primers. 2019;5(1):35.

4. Nirwan JS, Hasan SS, Babar ZU, Conway BR, Ghori MU. Global Prevalence and Risk Factors of Gastro-oesophageal Reflux Disease (GORD): Systematic Review with Meta-analysis. Sci Rep. 2020;10(1):5814.

5. Tack J, Pandolfino JE. Pathophysiology of Gastroesophageal Reflux Disease. Gastroenterology. 2018;154(2):277–88.

6. Zhang Z, Curran G, Altinok Dindar D, Wu Y, Wu H, Sharpton T, et al. Insights Into the Oral Microbiome and Barrett’s Esophagus Early Detection: A Narrative Review. Clin Transl Gastroenterol. 2021;12(9):e00390.

7. Pei Z, Bini EJ, Yang L, Zhou M, Francois F, Blaser MJ. Bacterial biota in the human distal esophagus. Proc Natl Acad Sci U S A. 2004;101(12):4250–5.

8. Peter S, Pendergraft A, VanDerPol W, Wilcox CM, Kyanam Kabir Baig KR, Morrow C, et al. Mucosa-Associated Microbiota in Barrett’s Esophagus, Dysplasia, and Esophageal Adenocarcinoma Differ Similarly Compared With Healthy Controls. Clin Transl Gastroenterol. 2020;11(8):e00199.

9. Yin J, Dong L, Zhao J, Wang H, Li J, Yu A, et al. Composition and consistence of the bacterial microbiome in upper, middle and lower esophagus before and after Lugol’s iodine staining in the esophagus cancer screening. Scand J Gastroenterol. 2020;55(12):1467–74.

10. Li Z, Dou L, Zhang Y, He S, Zhao D, Hao C, et al. Characterization of the Oral and Esophageal Microbiota in Esophageal Precancerous Lesions and Squamous Cell Carcinoma. Front Cell Infect Microbiol. 2021;11:714162.

11. Moreira C, Figueiredo C, Ferreira RM. The Role of the Microbiota in Esophageal Cancer. Cancers (Basel). 2023;15(9).

12. Yang L, Lu X, Nossa CW, Francois F, Peek RM, Pei Z. Inflammation and intestinal metaplasia of the distal esophagus are associated with alterations in the microbiome. Gastroenterology. 2009;137(2):588–97.

13. Corning B, Copland AP, Frye JW. The Esophageal Microbiome in Health and Disease. Curr Gastroenterol Rep. 2018;20(8):39.

14. Wu G, Xu T, Zhao N, Lam YY, Ding X, Wei D, et al. A core microbiome signature as an indicator of health. Cell. 2024;187(23):6550–65 e11.

15. Dempsey E, Corr SC. Lactobacillus spp. for Gastrointestinal Health: Current and Future Perspectives. Front Immunol. 2022;13:840245.

16. Matsuzaki T. Immunomodulation by treatment with Lactobacillus casei strain Shirota. Int J Food Microbiol. 1998;41(2):133–40.

17. Escamilla J, Lane MA, Maitin V. Cell-free supernatants from probiotic Lactobacillus casei and Lactobacillus rhamnosus GG decrease colon cancer cell invasion in vitro. Nutr Cancer. 2012;64(6):871–8.

18. Tiptiri-Kourpeti A, Spyridopoulou K, Santarmaki V, Aindelis G, Tompoulidou E, Lamprianidou EE, et al. Lactobacillus casei Exerts Anti-Proliferative Effects Accompanied by Apoptotic Cell Death and Up-Regulation of TRAIL in Colon Carcinoma Cells. PLoS One. 2016;11(2):e0147960.

19. Qian Y, Song JL, Sun P, Yi R, Liu H, Feng X, et al. Lactobacillus casei Strain Shirota Enhances the In Vitro Antiproliferative Effect of Geniposide in Human Oral Squamous Carcinoma HSC-3 Cells. Molecules. 2018;23(5).

20. Toi M, Hirota S, Tomotaki A, Sato N, Hozumi Y, Anan K, et al. Probiotic Beverage with Soy Isoflavone Consumption for Breast Cancer Prevention: A Case-control Study. Curr Nutr Food Sci. 2013;9(3):194–200.

21. Segata N, Izard J, Waldron L, Gevers D, Miropolsky L, Garrett WS, et al. Metagenomic biomarker discovery and explanation. Genome Biol. 2011;12(6):R60.

22. Lopetuso LR, Severgnini M, Pecere S, Ponziani FR, Boskoski I, Larghi A, et al. Esophageal microbiome signature in patients with Barrett’s esophagus and esophageal adenocarcinoma. PLoS One. 2020;15(5):e0231789.

23. Mozaffari Namin B, Soltan Dallal MM. Campylobacter Concisus and Its Effect on the Expression of CDX1 and COX2. Asian Pac J Cancer Prev. 2018;19(11):3211–6.

24. Mozaffari Namin B, Soltan Dallal MM, Ebrahimi Daryani N. The Effect of Campylobacter concisus on Expression of IL-18, TNF-alpha and p53 in Barrett’s Cell Lines. Jundishapur J Microbiol. 2015;8(12):e26393.

25. Blackett KL, Siddhi SS, Cleary S, Steed H, Miller MH, Macfarlane S, et al. Oesophageal bacterial biofilm changes in gastro-oesophageal reflux disease, Barrett’s and oesophageal carcinoma: association or causality? Aliment Pharmacol Ther. 2013;37(11):1084–92.

26. Macfarlane S, Furrie E, Macfarlane GT, Dillon JF. Microbial colonization of the upper gastrointestinal tract in patients with Barrett’s esophagus. Clin Infect Dis. 2007;45(1):29–38.

27. Greathouse KL, Stone JK, Vargas AJ, Choudhury A, Padgett RN, White JR, et al. Co-enrichment of cancer-associated bacterial taxa is correlated with immune cell infiltrates in esophageal tumor tissue. Sci Rep-Uk. 2024;14(1).

28. Ma JY, Piao XS, Mahfuz S, Long SF, Wang J. The interaction among gut microbes, the intestinal barrier and short chain fatty acids. Anim Nutr. 2022;9:159–74.

29. Horrocks V, King OG, Yip AYG, Marques IM, McDonald JAK. Role of the gut microbiota in nutrient competition and protection against intestinal pathogen colonization. Microbiol-Sgm. 2023;169(8).

30. Colombi E, Hill Y, Lines R, Sullivan JT, Kohlmeier MG, Christophersen CT, et al. Population genomics of Australian indigenous Mesorhizobium reveals diverse nonsymbiotic genospecies capable of nitrogen-fixing symbioses following horizontal gene transfer. Microb Genom. 2023;9(1).

31. Zhou J, Shrestha P, Qiu Z, Harman DG, Teoh WC, Al-Sohaily S, et al. Distinct Microbiota Dysbiosis in Patients with Non-Erosive Reflux Disease and Esophageal Adenocarcinoma. J Clin Med. 2020;9(7).

32. Satokari R. High Intake of Sugar and the Balance between Pro- and Anti-Inflammatory Gut Bacteria. Nutrients. 2020;12(5).

33. Elinav E, Strowig T, Kau AL, Henao-Mejia J, Thaiss CA, Booth CJ, et al. NLRP6 inflammasome regulates colonic microbial ecology and risk for colitis. Cell. 2011;145(5):745–57.

34. Scher JU, Sczesnak A, Longman RS, Segata N, Ubeda C, Bielski C, et al. Expansion of intestinal Prevotella copri correlates with enhanced susceptibility to arthritis. Elife. 2013;2:e01202.

35. Perdigon G, Maldonado Galdeano C, Valdez JC, Medici M. Interaction of lactic acid bacteria with the gut immune system. Eur J Clin Nutr. 2002;56 Suppl 4:S21–6.

36. Su SH, Mitani Y, Li T, Sachdeva U, Flashner S, Klein-Szanto A, et al. Lactate Suppresses Growth of Esophageal Adenocarcinoma Patient-Derived Organoids through Alterations in Tumor NADH/NAD+ Redox State. Biomolecules. 2024;14(9).

37. Li D, He R, Hou G, Ming W, Fan T, Chen L, et al. Characterization of the Esophageal Microbiota and Prediction of the Metabolic Pathways Involved in Esophageal Cancer. Front Cell Infect Microbiol. 2020;10:268.

38. Tiwari A, Ika Krisnawati D, Susilowati E, Mutalik C, Kuo TR. Next-Generation Probiotics and Chronic Diseases: A Review of Current Research and Future Directions. J Agric Food Chem. 2024;72(50):27679–700.

