## Supplementary material for "*Lactobacillus casei* Shirota intervention modulates the esophageal microbiome composition in Barrett’s esophagus": Supplemetnal files

* Corresponding Author:

Dr. Annemarie Boleij

#### Content:

1. Supplemental methods
2. Supplemental figures
3. Supplemental tables

### 1. Supplemental methods

#### Sampling procedures

At baseline and after 28 ± 3 days of LcS intake, participants underwent upper endoscopy with biopsy sampling. All study endoscopies were performed by one experienced endoscopist (PDS). The length of BE was measured using the Prague classification.(1) At both endoscopies, four-quadrant biopsies were taken every 2 cm according to the Seattle protocol, with additional targeted biopsy of any macroscopic abnormalities.(2) Biopsies were fixed in buffered formalin for histopathological examination by a local gastrointestinal pathologists.

Additional biopsy samples from normal squamous epithelium (NSE, n = 4) and metaplastic columnar epithelium (MCE, n = 4) were taken for study purposes during each endoscopy. To minimize cross-contamination, samples of MCE were collected first before NSE, and biopsy forceps were washed in sterile water between samples. MCE biopsies were obtained at least 1 cm above the gastroesophageal junction and biopsies of NSE at least 3 cm above the squamocolumnar junction. For each patient, 2 NSE and 2 MCE biopsies were fixed in Guanidine Hydrochloride for DNA and RNA isolation for LcS-analysis and microbial qPCRs and 1 NSE and 1 MCE biopsy were fresh frozen in liquid nitrogen and immediately transferred to -80°C for 16s rDNA analysis of the esophageal microbiome. Two study biopsies 1 NSE and 1 MCE were fixated in Methacarn (30% chloroform, 10% acetic acid, 60% methanol) for *in situ* bacterial analysis. Study biopsies were scored by a trained pathologist for the presence of Barrett epithelium (metaplastic epithelium with intestinal metaplasia), gastric esophageal junction, gastric cardia or squamous epithelium on HE stained sections. An inflammation score from no inflammation, minimal, moderate or severe chronic inflammation was assigned. Researchers who performed subsequent processes were blinded to clinical information. For analysis performed on each biopsy see **Supplemental table S1**.

#### Tissue processing

Histology and Fluorescence in situ hybridization (FISH): Tissue sections of 4 µm were used for Hematoxylin/Eosin (HE) staining and FISH. Stained slides were scanned at 20x magnification using the Pannoramic 1000 scanner for HE slides and Pannoramic Midi fluorescence scanner (3D Histech) for FISH with the following exposure times: DAPI 20 ms , FITC 40 ms, TRITC 38 ms, and Cy5 38 ms. HE slides were scored by a pathologist to verify presence of NSE and MCE in the obtained biopsies from each location. FISH was performed to detect bacteria in NSE and MCE tissue samples. Tissue sections were deparaffinized in xylene and rehydrated through a series of ethanol washes. FISH probes (EUB338-cy5 and non-EUB338-cy5) were diluted in hybridization buffer (0.9 M NaCl, 20 mM Tris-HCl pH 8.0, 0.01% sodium dodecyl sulfate) and added to airdried sections. For the probe mixture to detect Bacteroidetes (Alexa488), Firmicutes (cy3), and Gammaproteobacteria (cy5) 20% formamide was added to the hybridization buffer to optimize hybridization of the probes. All probes were tested on protoblocks of bacterial strains Streptococcus gallolyticus, Escherichia coli and Bacteroides fragilis, and colon eptihelial tissue containing a biofilm to test specificity of the probes (**Supplemental figure S1**). Sections were incubated in humidified box overnight at 46°C and washed 3 times for 5 minutes in washing buffer at 48°C (0.9M NaCl, 20mM Tris-Cl pH 8.0), followed by 1 minute iced H_2_O and 1 minute in PBS. Slide mounting was performed with Prolong Gold Antifade medium with DAPI (Thermo Fisher Scientific, P36931). The EUB338 probe was used for all slides and for the positive control and the non-EUB probe was used to verify positive signals and exclude unspecific probe binding. FISH was carried out with oligonucleotide probes targeting bacterial 16S rRNA as listed in **Supplemental table S2.**

DNA isolation: Endoscopic biopsies collected in liquid nitrogen were utilized to extract DNA from bacteria present in MCE and NSE. DNA extraction from mucosal biopsies was performed according to a previously optimized protocol(3). Briefly, frozen biopsies were placed in PBS , thawed and mixed for 5 minutes. Loosely attached bacteria were collected by removing the PBS from the biopsy. This biopsy wash was saved on ice for later extraction. Firmly attached bacteria were recovered by proteinase K tissue digestion. The digested biopsy and PBS wash were combined; human DNA was digested by a combination of mild lysis with saponin (preserving bacterial cells) and DNAse digestion. Intact bacteria were collected by centrifugation at 10,000g for 10 minutes at 4°C. The bacterial pellet was suspended in Power bead solution containing mutanolysin from *Streptomyces globisporus* (ATCC 21553) and incubated at 37°C for 60 minutes. Subsequent DNA extraction was performed according to the DNeasy Powerlyzer Powersoil kit (Qiagen), including bacterial lyses with 0.1 mm glass beads using the Fisherbrand Bead Mill 4, two times at maximum speed for 30 sec with 30 sec rest on ice in between. The total DNA yield was quantified using the Qubit 5.0 Fluorometer Broad Range DNA assay (Cat #Q32853, Life Technologies, USA), following the manufacturer's guidelines. DNA quality was assessed by DNA electrophoresis in 1% agarose.

#### Scoring of HE & FISH slides

Each biopsy from NSE and MCE regions was evaluated by apathologist specialized in upper gastointestinal pathology (R.S. van der Post) to confirm that NSE biopsies were from squamous epithelium and to confirm that the MCE regions contained columnar epithelium representative of BE. Each FISH image was compared to a non-EUB stained control to determine specificity of the signals. First the length of the apical region in the biopsy was determined to control for the size of the biopsies represented in the slices that were taken. Next all positive signals along the length of the epithelium were counted and divided by the length of the epithelium to generate number of bacteria per mm tissue. The same method was perform for each probe (LGC-mix, CFB, Gam42 and Eub338). All FISH slides were scored by two observers and a third for confirmation of the results.

#### Quantitative PCR analysis for Gram-positive /Gram-negative ratio and LcS

To determine the ratio of Gram-positive (G+) and Gram-negative (G-) bacteria one set of primers and two probes were used. Primers PLK1 (5’-TACGGGAGGCAGCAGT-3’) and PLK2 (5’-TATTACCGCGGCTGCT-3’) targeting bacterial 16S rRNA were selected for amplification (4). Internal hybridization probes G+ (5’-*FAM-*CTAACCAGAAAGCCACGGCTAACTACGTG–*OQA*-3’) and G−(5’-*HEX*–TTACCCGCAGAATAAGCACCGGCTAAC–*BHQ1*-3’) were used for detecting the amplified template(5). For *Lactobacillus casei* strain Shirota (*LcS)* detection, the *LcS-*specific primer set pLcS with primers pLcS-57F (5’-CTCAAAGCCGTGACGGTC-3’) and pLcS-597R (5’-CACTAGGATTATTAGCACCACGT-3’) were selected for amplification (6). The internal hybridization probe LcS (5’-*FAM*-CCTCTTGGGGAACCAGTGCAGCAG-3’) was designed to detect the amplified DNA template of the pLcS primer set. The primer probe combination was validated using LcS DNA and a set of pooled DNA from 17 different bacteria as negative control (*Acinetobacter baumannii, Enterococcus faecalis, Enterococcus faecium, Citrobacter freundii, Enterobacter cloacae, Staphylococcus aureus, Escherichia coli, Staphylococcus epidermidis, Klebsiella oxytoca, Klebsiella pneumoniae, Streptococcus agalactiae, Streptococcus pneumoniae, Streptococcus pyogenes, Proteus mirabilis, Morganella morganii, Pseudomonas aeruginosa, Staphylococcus warneri)*. The qPCR reactions were performed using the QX200 Droplet Digital PCR system (Bio-Rad, CA, USA) in a final volume of 22 µl ddPCR mastermix. Droplets were generated using the QX200 Droplet Generator. The generated droplet suspensions were transferred into a 96‐wells plate and amplified by using a C1000 Thermal Cycler (Bio‐Rad). The G+/G- PCR cycling conditions consisted of 95°C for 10 min, 40 cycles of 95°C for 30 s and 61°C for 1 min, and a final step at 98°C for 10 min. The LcS quantification conditions consisted of 95°C for 10 min, 40 cycles of 95°C for 30 s, 63°C for 1 min and 72°C for 30 s, and a final step at 98°C for 10 min. After amplification, the samples were measured with the QX200 Droplet Reader (Bio‐Rad) and the results were processed and shown with QuantaSoft software version 1.7.4 (Bio‐Rad). G+/G- -ratio and LcS copies/μl were automatically determined by the QuantaSoft software after manually setting the thresholds above the negative (signal‐arm) droplets.

#### Microbiome analysis

Microbiota profiling for taxonomic classification by amplicon sequencing was carried out by Novogene Co. To this, 250 bp (paired-end) of sequence from 16S rRNA genes of distinct regions (16SV4/16SV3/16SV3-V4/16SV4-V5) were amplified used specific primers (e.g. 16S V4: 515F-806R, 18S V4: 528F-706R, 18S V9: 1380F-1510R, et. al ) with barcode. PCR reactions were carried out with Phusion® High-Fidelity PCR Master Mix (New England Biolabs). PCR products were mixed at equal density ratios, and mixed PCR products were purified with Qiagen Gel Extraction Kit (Qiagen, Germany). Sequencing libraries generated with NEBNext® UltraTM DNA Library Prep Kit for Illumina and quantified via Qubit and Q-PCR. Sequencing was performed using the Illumina Novaseq 6000 platform.

**Processing of sequencing data** - Paired-end reads were assigned to samples based on their unique barcodes and truncated by cutting off the barcode and primer sequences. Paired-end reads were merged using FLASH (V1.2.7) (7) (<http://ccb.jhu.edu/software/FLASH/>), a fast and accurate analysis tool, which was designed to merge paired-end reads when at least some of the reads overlap the read generated from the opposite end of the same DNA fragment, and the splicing sequences were called raw tags. Quality filtering on the raw tags were performed under specific filtering conditions to obtain the high-quality clean tags (8) according to the Qiime (V1.7.0) workflow (9) (<http://qiime.org/scripts/split_libraries_fastq.html>) quality controlled process.

Tags were compared to the reference database (Gold database) using the UCHIME algorithm (10) (<http://www.drive5.com/usearch/manual/uchime_algo.html>) to detect chimera sequences (<https://drive5.com/usearch/manual/chimeras.html>) and remove chimeras (11) to obtain the final Effective Tags.

**OTU cluster and Taxonomic annotation** - Sequence analysis was performed by Uparse software (Uparse v7.0.1001, <http://drive5.com/uparse/>) (12) using the effective tags. Sequences with ≥97% similarity were assigned to the same OTUs. Representative sequence for each OTU was screened for further annotation. For each representative sequence, Mothur software (13) was used to compare reads against the SSUrRNA database of SILVA Database ([http://www.arb-silva.de/)](http://www.arb-silva.de/)%20%5b26) for species annotation at each taxonomic rank (Threshold:0.8~1; kingdom, phylum, class, order, family, genus, species) (14).

To obtain the phylogenetic relationship of all OTUs representative sequences, the MUSCLE (Version 3.8.31, <http://www.drive5.com/muscle/>) aligner (15) was used for fast comparison of sequences. OTUs abundances were normalized using a standard of sequence number corresponding to the sample with the least sequences. Subsequent analysis of alpha diversity and beta diversity were all performed basing on this output normalized data.

**Alpha Diversity** - Alpha diversity was applied to analyze complexity of biodiversity for a sample through 6 indices, including Observed-species, Chao1, Shannon, Simpson, ACE, Good-coverage. All indices in samples were calculated with QIIME (Version 1.7.0) and displayed with R software (Version 2.15.3). The following Alpha Diversity Indices were calculated:

Community richness indices: Chao - the Chao1 estimator (<http://scikit-bio.org/docs/latest/generated/skbio.diversity.alpha.chao1.html#skbio.diversity.alpha.chao1>);

ACE - the ACE estimator (<http://scikit-bio.org/docs/latest/generated/skbio.diversity.alpha.ace.html#skbio.diversity.alpha.ace>);

Community diversity indices:

Shannon - the Shannon index (<http://scikit-bio.org/docs/latest/generated/skbio.diversity.alpha.shannon.html#skbio.diversity.alpha.shannon>);"

Simpson - the Simpson index (<http://scikit-bio.org/docs/latest/generated/skbio.diversity.alpha.simpson.html#skbio.diversity.alpha.simpson>);

The index of sequencing depth:

Coverage - the Good’s coverage (<http://scikit-bio.org/docs/latest/generated/skbio.diversity.alpha.goods_coverage.html#skbio.diversity.alpha.goods_coverage>);

The index of phylogenetic diversity:

PD_whole_tree - PD_whole_tree index (<http://scikit-bio.org/docs/latest/generated/skbio.diversity.alpha.faith_pd.html?highlight=pd#skbio.diversity.alpha.faith_pd>)

**Beta Diversity -** Beta diversity analysis was used to evaluate differences of samples in species complexity, Beta diversity on both weighted and unweighted unifrac were calculated by QIIME software (Version 1.7.0). Cluster analysis was preceded by principal component analysis (PCA), which was applied to reduce the dimension of the original variables using the FactoMineR package and ggplot2 package in R software (Version 2.15.3). Principal Coordinate Analysis (PCoA) was performed to get principal coordinates and visualize from complex, multidimensional data. A distance matrix of weighted or unweighted unifrac among samples obtained before was transformed to a new set of orthogonal axes, by which the maximum variation factor is demonstrated by first principal coordinate, and the second maximum one by the second principal coordinate, and so on. PCoA analysis was displayed by WGCNA package, stat packages and ggplot2 package in R software (Version 2.15.3). Unweighted Pair-group Method with Arithmetic Means (UPGMA) Clustering was performed as a type of hierarchical clustering method to interpret the distance matrix using average linkage and was conducted by QIIME software (Version 1.7.0).

Metastat was calculated by R software. P-value was calculated by method of permutation test while q-value was calculated by method of Benjamini and Hochberg False Discovery Rate (16). Anosim, MRPP and Adonis were performed by R software (Vegan package: anosim function, mrpp function and adonis function). AMOVA was calculated by mothur using amova function. T_test and drawing were conducted by R software.

The raw sequence files were deposited in the European nulceotide archive (ENA) under accession number PRJEB89339.

Post-hoc pairwise comparisons of alpha-diversity for all group comparisons at the genus level were assessed using the Shannon index with Mann-Whitney or Kruskal-Wallis tests. The multi-testing adjustment is based on the Benjamini-Hochberg procedure (FDR). The differences in microbial composition and diversity between experimental groups were evaluated using the Bray–Curtis dissimilarity index. Permutational multivariate analysis of variance (PERMANOVA) was performed on the genera-level abundance profiles of the samples to assess the effects of location sites and interventions. The PERMANOVA and all p values were adjusted for false discovery rate (FDR) using the Benjamini–Hochberg procedure. Principal coordinate analyses (PCoA), Principal component analysis (PCA), partial-, and redundancy analysis (RDA) were visualized and assessed using the Canoco 5.10 software suite with 1000 permutations to measure significance and where applicable, corrected for covariates (histological type and patients; LcS intervention and patients). Where necessary, relative abundance values were log-transformed (Y’ = log(Y + 1000)). The RDA plot figures show the top 10 bacterial species with the highest RDA values.

Differential abundance of specific microbiota taxa in groups (tissue type, before or after LcS intervention) were also characterized using the Linear Discriminant Analysis (LDA) effect size (LEfSe) procedure (see main text), employed to classify and reduce dimensionality by identifying the optimal linear combination of taxa for the experimental variable (e.g., esophageal regions and intervention). LEfSe identifies the characteristics (organisms, clades, operational taxonomic units, genes, or functions) most likely to elucidate differences across classes by integrating traditional statistical significance tests with further assessments that account for biological consistency and effect relevance. The Kruskal-Wallis rank-sum test and effect size measurements were applied to detect differentially abundant taxa between the two groups in each comparison.

#### Statistical analysis

FISH analysis: A Wilcoxon signed-rank test was used to determine the significance of differences in Gram-positive/Gram-negative -ratio between baseline (day=0) and after LcS intervention (day=29) within each sample type group and to evaluate the difference in relative abundance of specific bacterial taxa per mm tissue between baseline and after intervention. A paired Wilcoxon signed-rank test was used to determine the difference in relative abundance of specific bacterial taxa between NSE and MCE before, and after intervention. The p-value <0,05 cutoff was used to determine statistical significance. All data were analyzed in GraphPad Prism 10. v1.2 (Dotmatics).

### 2. Supplemental Figures

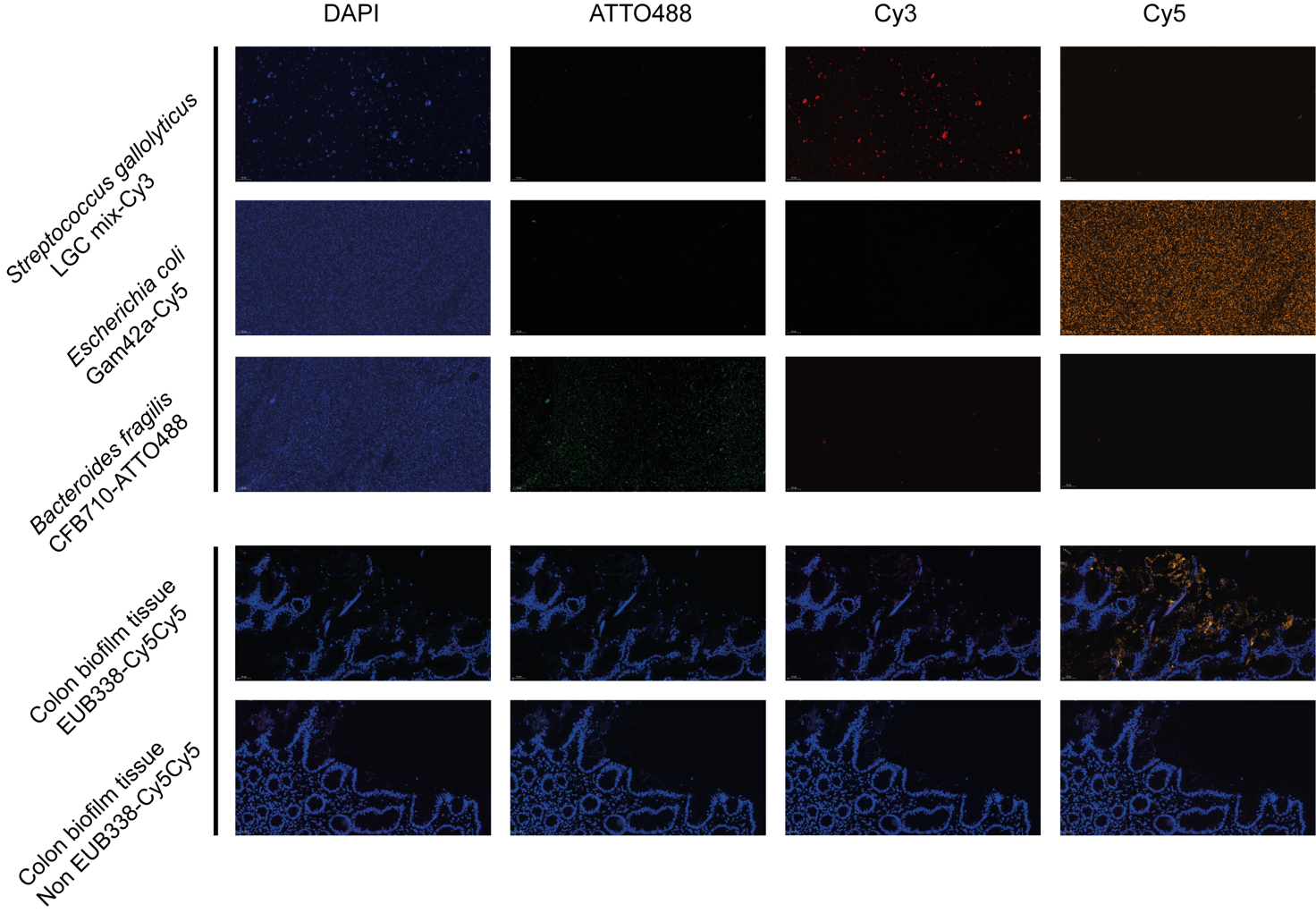

**A**

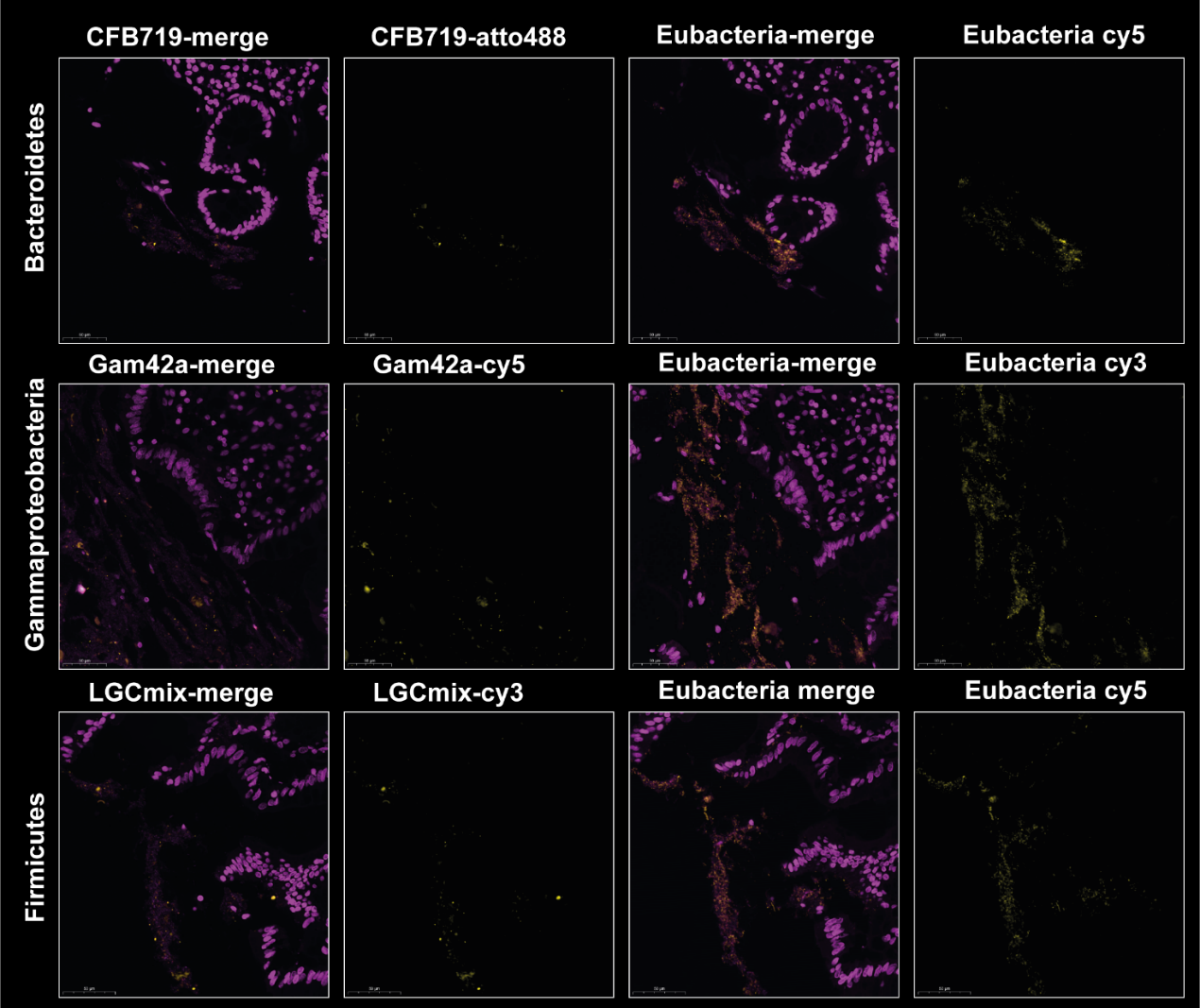

**B**

**Supplemental Figure S1.** **A) Representative FISH images of control bacteria and colon tissue**. Validated FISH probes for *in situ* detection of Firmicutes (LGC-mix-cy3 (red)), Bacteroidetes (CFB719-ATTO488(green)) and Gammaproteobacteria (Gam42a (orange)) were tested on agar embedded bacteria in 4µm slices. These validated probes were selected from Probe-base (17) and tested on *Streptococcus gallolyticus* representing Firmicutes, *Escherichia coli* representing Gammaproteobacteria and *Bacteroides fragilis* representing Bacteroidetes. All probes worked properly and for each staining a non-EUB338 control was taken along to confirm specificity of the signals. The eub338cy5cy5 probe (orange) was used to detect Eubacteria. A colon biopsy with biofilm was used as a positive control in the experiments. DAPI (blue) stains the nuclei and bacterial DNA. **B)** **FISH images of control colon tissue**. FISH probes for Firmicutes (LGC-mix-cy3), Bacteroidetes (CFB719-ATTO488) and Gammaproteobacteria (Gam42a) were tested on a control colon biopsy with a biofilm to test the feasibility of the probes on human tissue. All specific probes were simultaneously stained with EUB338 detecting Eubacteria labeled with cy5 or cy3 depending on the label of the specific probe. Bacteroidetes, Gammaproteobacteria and Firmicutes were detected in the colon biofilm. The DNA is stained with DAPI and visible in the merged image in purple. The bacterial signal in each image is depicted in yellow.

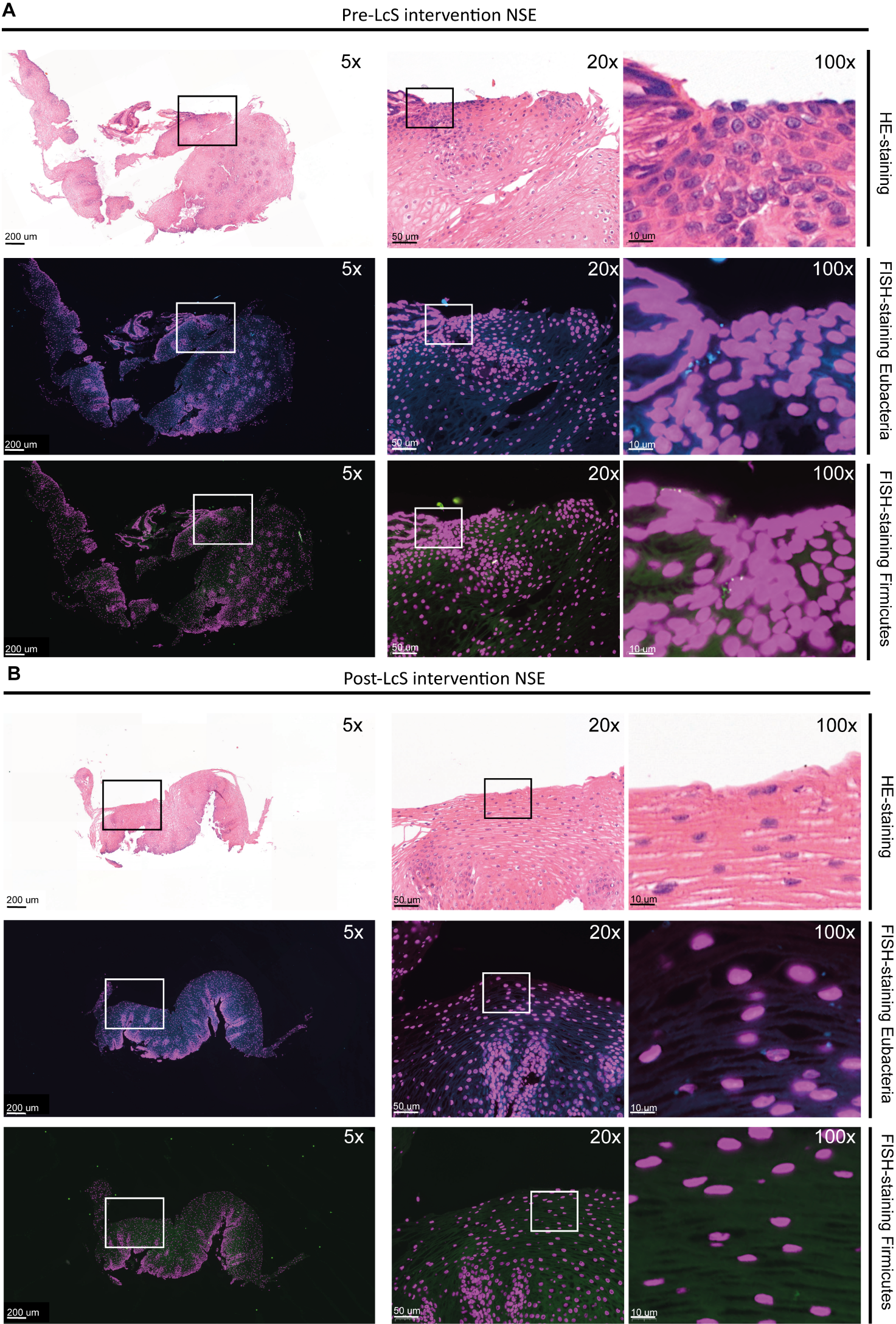

#### Supplemental figure S2. *In situ* bacterial detection of Eubacteria and Firmicutes in NSE biopsies (A) pre- and (B) post-LcS intervention. Eubacteria are depicted in cyan, Firmicutes are depicted in green and DAPI is depicted in Magenta.

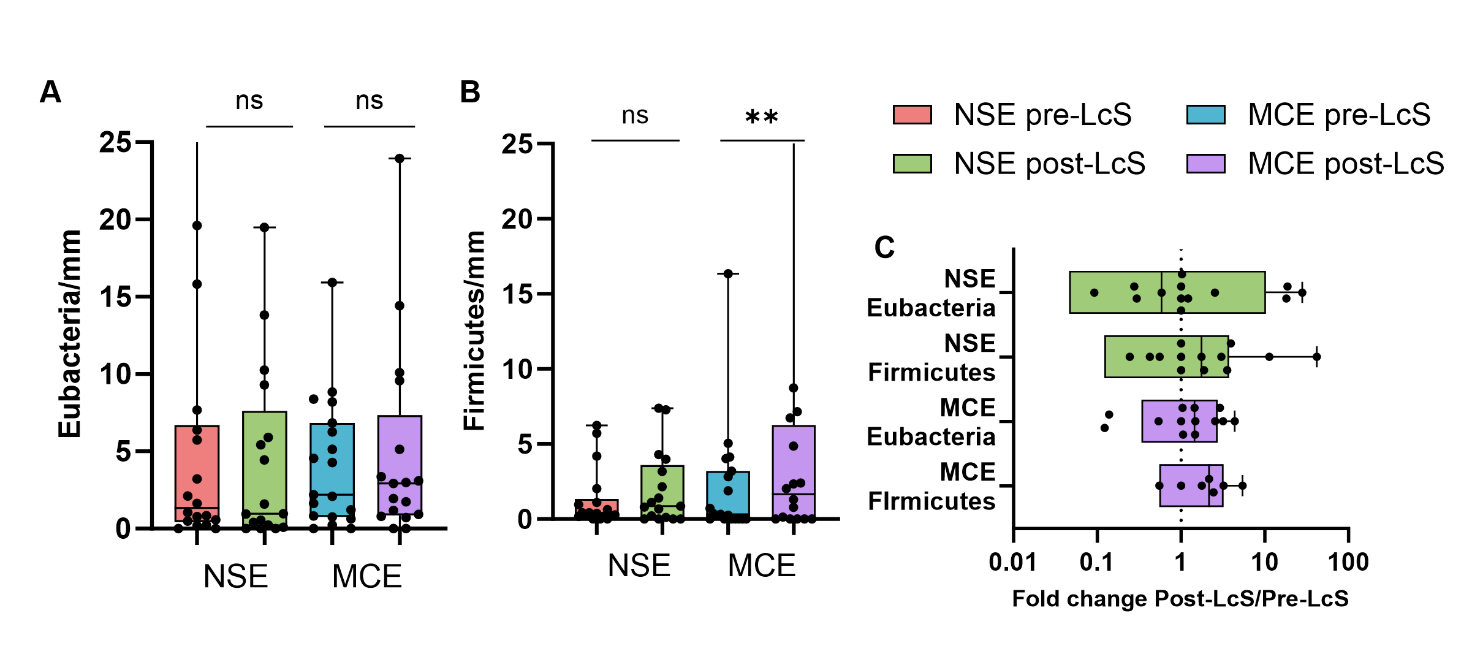

**Supplemental figure S3:** A) Eubacteria signals detected per mm apical region in NSE and MCE biopsies. No statistically significant difference was observed in the number of Eubacteria signals per mm in both MCE (median 2.19, and 2.93), and NSE biopsies (median 1.35 and 0.96) pre- and post-LcS, respectively. B) A significant increase in Firmicutes signals post-LcS was observed per mm apical region for MCE biopsies (median 0.32 vs 1.67; ** p < 0,01 Wilcoxon Signed Rank test), but no significant increase in Firmicutes post-LCS was observed in NSE biopsies (median 0.4 vs 0.86). C) the ratio of bacteria per mm tissue post- versus pre-LcS is depicted on the x-axis, a ratio of 1 means no difference post- versus pre-LcS, a positive ratio means an increase post-LcS, a fold change of >2 was observed for Firmicutes in MCE biopsies in 5 out of 7 paired biopsies.

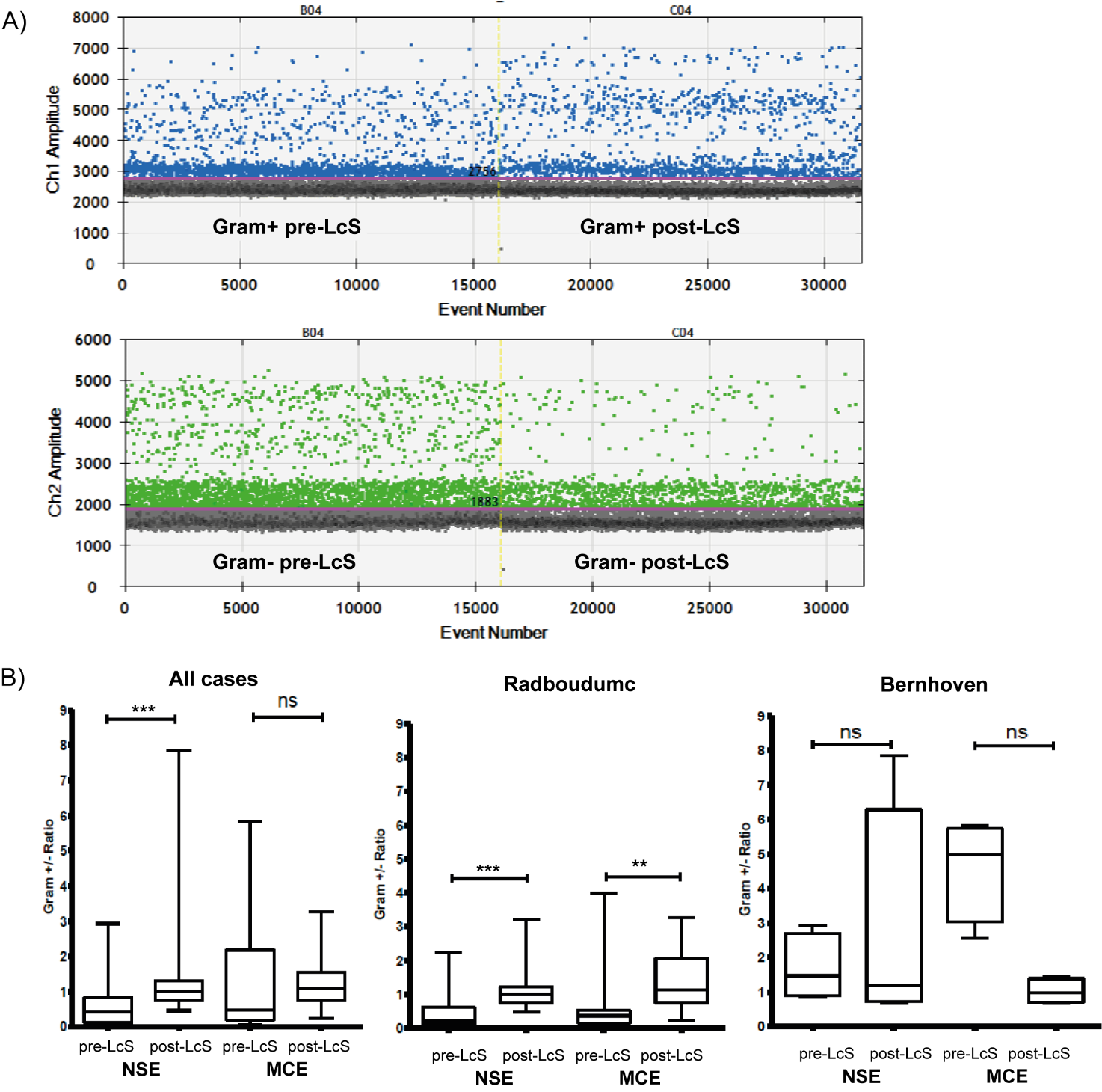

**Supplemental Figure S4.**

A) Example of Gram+ and Gram- bacteria by ddPCR detection in MCE biopsies pre- and post-LcS intervention. A notable increase in Gram-positive bacteria and decrease in Gram-negative bacteria is observed. Blue dots on channel 1 (y-axis) represent Gram+ bacteria; Green dots on channel 2 (y-axis) represents Gram- bacteria (Gram-); Event number on the x axis showcase the total droplet count analyzed. Amplitude represented by the y-axis accounts for total signal per droplet. Droplets above the threshold line (purple line) were determined as positive result. B) Changes in Gram+ /Gram- ratio (y axis) of all patients (n=20), only Radboudumc patients (n=16) and only Bernhoven patients (n=4) pre- and post-LcS intervention according to 16S rRNA ddPCR analyses. A significant increase of 0.42 to 1.02 in Gram+ /Gram- ratio for NSE was observed (*** p=0.003, Wilcoxon signed-rank test), and a non-significant increase of 0.48 to 1.11 for MCE biopsies. Separate analysis of the Radboudumc patients (n=16) showed a significant increase from 0.21 to 1.02 (*** p=0.004) for NSE and 0.36 to1.12 (** p=0.008) for MCE post-LcS intervention. The Bernhoven patients (n=4) subgroup was too small to test for statistical significance in separate analysis.

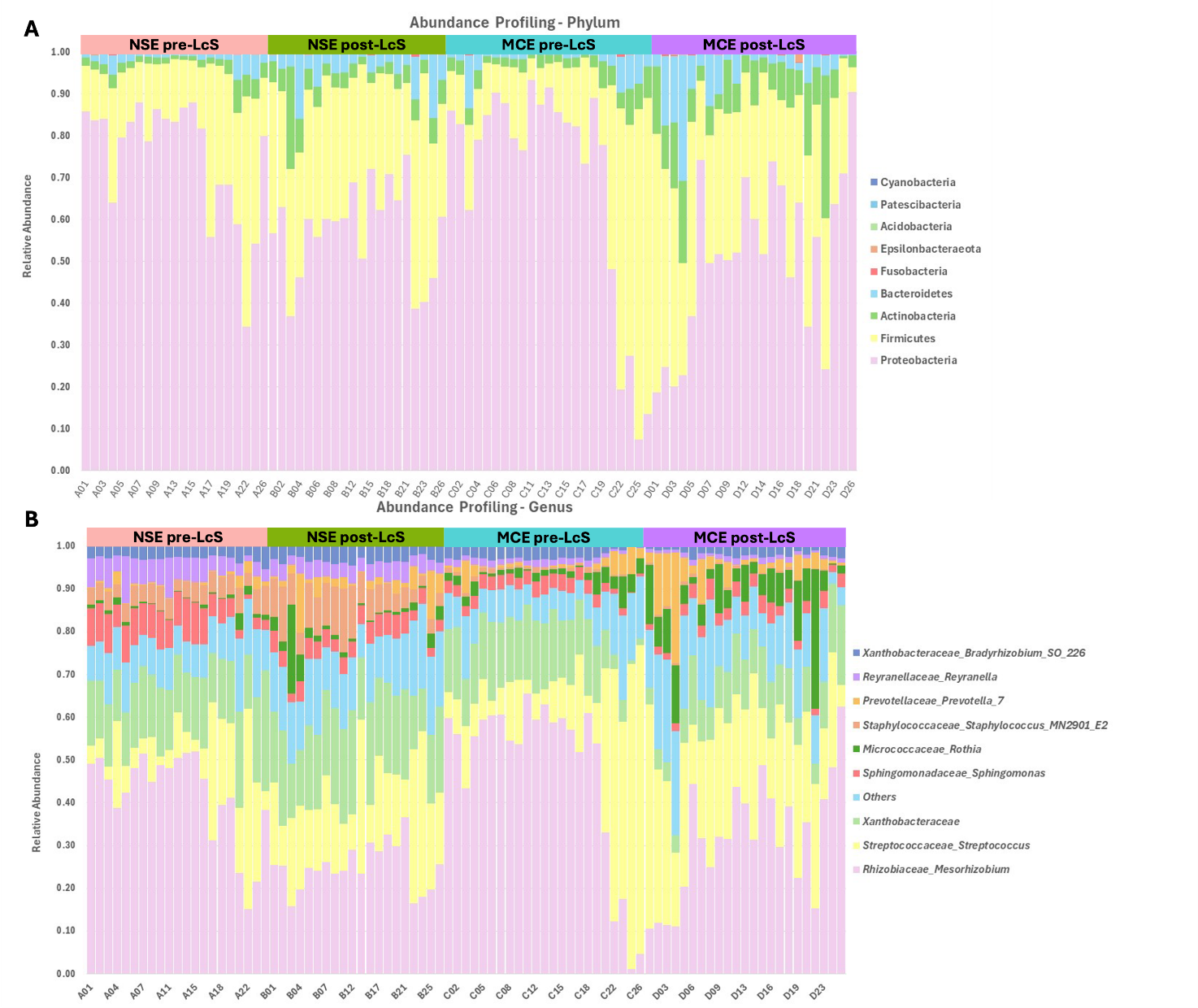
**Supplemental Figure S5.** **Relative abundance of bacterial taxa at Phylum (A) and Genus (B) level in NSE and MCE samples pre- and post-LcS.** Phylogenetic profiles were more similar within individuals than between the two sampled anatomical locations with either normal healthy (squamous (NSE)) or BE (columnar) morphology (MCE). Microbial profile of top ten taxa at phylum and genus level in patients affected by BE in two different esophageal locations (NSE (orange) and MCE (green)) before (pre-LcS) and after intervention (post-LcS) with LcS is shown at phylum (A) and genus (B) level. Individuals are grouped for the four experimental categories.

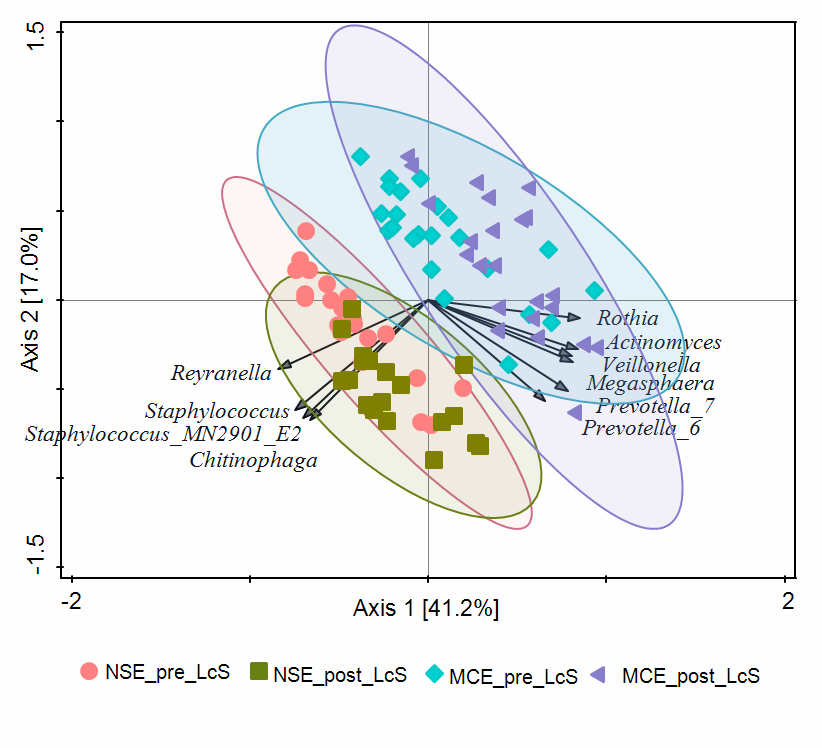
**Supplemental Figure S6. Principal component analysis of microbiota composition.**

Principal component analysis (PCA) visualizes differences in microbiota composition at genus level associated with sampled esophageal regions and pre- or post-LcS intervention sampling time points. PCA was performed at the genus level using individual samples, which are represented by symbols for each experimental group. The top 10 genera with the highest contributions to the principal components (PC) are plotted as arrows. Ellipses represent 95% confidence intervals for the groupings. The percentages along the axes indicate the proportion of variation explained by the PC; both axes explain nearly 60% of the total variation in the data.

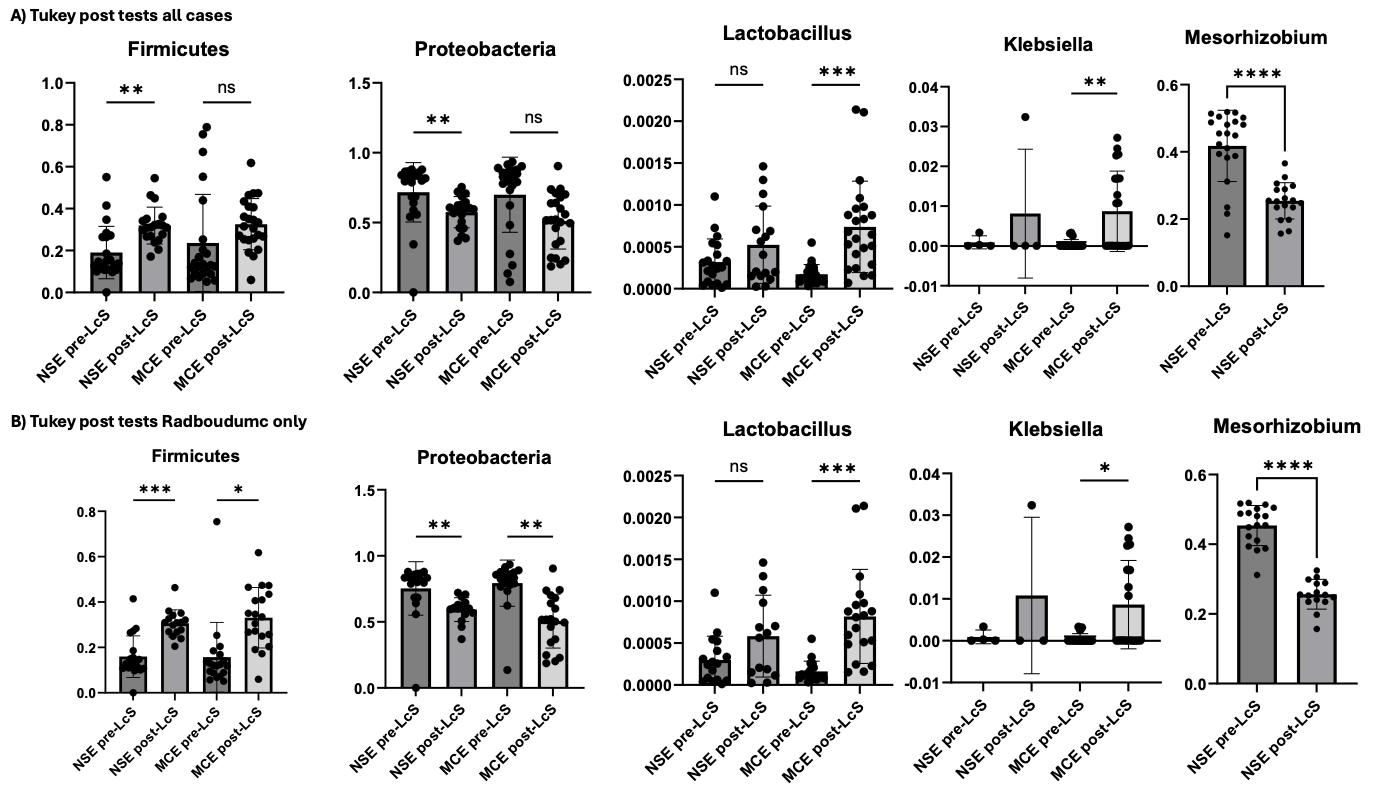

**Supplemental Figure S7. Tukey's post hoc test of relative abundance of taxa pre- and post LcS in NSE and MCE biopsies.** A) Relative abundance plots of all cases (including Bernhoven hospital; n=23), and B) of patients from the Radboudumc hospital only (n=19). Relative abundance plots are plotted foreach group separately (NSE pre-LcS, NSE post-LcS, MCE pre-LcS and MCE post-LcS). *p<0.05, **p<0.01, ***p<0.005, **** p<0.001.

### Supplemental Tables

#### Supplemental Table S1. Overview analysis performed on each case and sample

| **Case** | **Group** | **Inclusion** | **Hospital** | **Clinical Location** | **LcS** | **Clinical histology (FFPE)** | **Study histology (MCPE)** | **FISH (MCPE)** | **16s seq (frozen)** | **ddPCR (GH)** |
| --- | --- | --- | --- | --- | --- | --- | --- | --- | --- | --- |
| 1 | A | Yes | Radboudumc | NSE | pre_LcS | Yes | No | No | Yes | No |
| 2 | A | Yes | Radboudumc | NSE | pre_LcS | Yes | No | No | Yes | Yes |
| 3 | A | Yes | Radboudumc | NSE | pre_LcS | Yes | No | No | Yes | Yes |
| 4 | A | Yes | Radboudumc | NSE | pre_LcS | Yes | No | No | Yes | Yes |
| 5 | A | Yes | Radboudumc | NSE | pre_LcS | Yes | Yes | Yes | Yes | Yes |
| 6 | A | Yes | Radboudumc | NSE | pre_LcS | Yes | Yes | Yes | Yes | Yes |
| 7 | A | Yes | Radboudumc | NSE | pre_LcS | Yes | Yes | Yes | Yes | Yes |
| 8 | A | Yes | Radboudumc | NSE | pre_LcS | Yes | Yes | Yes | Yes | Yes |
| 9 | A | Yes | Radboudumc | NSE | pre_LcS | Yes | Yes | Yes | Yes | No |
| 10 | A | No | Radboudumc | NSE | pre_LcS | Yes | No | No | No | No |
| 11 | A | Yes | Radboudumc | NSE | pre_LcS | Yes | Yes | Yes | Yes | Yes |
| 12 | A | yes | Radboudumc | NSE | pre-LcS | Yes | Yes | Yes | No | No |
| 13 | A | Yes | Radboudumc | NSE | pre_LcS | Yes | Yes | Yes | Yes | Yes |
| 14 | A | Yes | Radboudumc | NSE | pre_LcS | Yes | Yes | Yes | Yes | Yes |
| 15 | A | Yes | Radboudumc | NSE | pre_LcS | Yes | Yes | Yes | Yes | Yes |
| 16 | A | Yes | Radboudumc | NSE | pre_LcS | Yes | Yes | Yes | Yes | Yes |
| 17 | A | Yes | Radboudumc | NSE | pre_LcS | Yes | Yes | Yes | Yes | Yes |
| 18 | A | Yes | Radboudumc | NSE | pre_LcS | Yes | No | No | Yes | Yes |
| 19 | A | Yes | Radboudumc | NSE | pre_LcS | Yes | Yes | Yes | Yes | Yes |
| 21 | A | Yes | Bernhoven | NSE | pre_LcS | Yes | Yes | Yes | Yes | Yes |
| 22 | A | Yes | Bernhoven | NSE | pre_LcS | Yes | Yes | Yes | Yes | Yes |
| 23 | A | Yes | Bernhoven | NSE | pre_LcS | Yes | Yes | Yes | No | Yes |
| 24 | A | No | Bernhoven | NSE | pre_LcS | Yes | No | No | No | No |
| 25 | A | Yes | Bernhoven | NSE | pre_LcS | Yes | Yes | Yes | Yes | Yes |
| 26 | A | Yes | Radboudumc | NSE | pre_LcS | Yes | Yes | Yes | Yes | Yes |
| 1 | B | Yes | Radboudumc | NSE | post_LcS | No | No | No | Yes | No |
| 2 | B | Yes | Radboudumc | NSE | post_LcS | No | No | No | Yes | Yes |
| 3 | B | Yes | Radboudumc | NSE | post_LcS | No | No | No | Yes | Yes |
| 4 | B | Yes | Radboudumc | NSE | post_LcS | No | No | No | Yes | Yes |
| 5 | B | Yes | Radboudumc | NSE | post_LcS | No | Yes | Yes | Yes | Yes |
| 6 | B | Yes | Radboudumc | NSE | post_LcS | No | Yes | Yes | Yes | Yes |
| 7 | B | Yes | Radboudumc | NSE | post_LcS | No | Yes | Yes | Yes | Yes |
| 8 | B | Yes | Radboudumc | NSE | post_LcS | No | Yes | Yes | Yes | Yes |
| 9 | B | Yes | Radboudumc | NSE | post_LcS | No | Yes | Yes | No | No |
| 10 | B | No | Radboudumc | NSE | post_LcS | No | No | No | No | No |
| 11 | B | Yes | Radboudumc | NSE | post_LcS | No | Yes | Yes | Yes | Yes |
| 12 | B | Yes | Radboudumc | NSE | post_LcS | No | Yes | Yes | Yes | No |
| 13 | B | Yes | Radboudumc | NSE | post_LcS | No | Yes | Yes | Yes | Yes |
| 14 | B | Yes | Radboudumc | NSE | post_LcS | No | Yes | Yes | No | Yes |
| 15 | B | Yes | Radboudumc | NSE | post_LcS | No | Yes | No | Yes | Yes |
| 16 | B | Yes | Radboudumc | NSE | post_LcS | No | Yes | Yes | No | Yes |
| 17 | B | Yes | Radboudumc | NSE | post_LcS | No | No | No | Yes | Yes |
| 18 | B | Yes | Radboudumc | NSE | post_LcS | No | Yes | Yes | Yes | Yes |
| 19 | B | Yes | Radboudumc | NSE | post_LcS | No | Yes | Yes | Yes | Yes |
| 21 | B | Yes | Bernhoven | NSE | post_LcS | No | Yes | Yes | Yes | Yes |
| 22 | B | Yes | Bernhoven | NSE | post_LcS | No | Yes | Yes | Yes | Yes |
| 23 | B | Yes | Bernhoven | NSE | post_LcS | No | Yes | Yes | Yes | Yes |
| 24 | B | No | Bernhoven | NSE | post_LcS | No | No | No | No | No |
| 25 | B | Yes | Bernhoven | NSE | post_LcS | No | Yes | Yes | Yes | Yes |
| 26 | B | Yes | Radboudumc | NSE | post_LcS | No | Yes | Yes | Yes | Yes |
| 1 | C | Yes | Radboudumc | MCE | pre_LcS | Yes | No | No | Yes | No |
| 2 | C | Yes | Radboudumc | MCE | pre_LcS | Yes | No | No | Yes | Yes |
| 3 | C | Yes | Radboudumc | MCE | pre_LcS | Yes | No | No | Yes | Yes |
| 4 | C | Yes | Radboudumc | MCE | pre_LcS | Yes | No | No | Yes | Yes |
| 5 | C | Yes | Radboudumc | MCE | pre_LcS | Yes | Yes | Yes | Yes | Yes |
| 6 | C | Yes | Radboudumc | MCE | pre_LcS | Yes | Yes | Yes | Yes | Yes |
| 7 | C | Yes | Radboudumc | MCE | pre_LcS | Yes | Yes | Yes | Yes | Yes |
| 8 | C | Yes | Radboudumc | MCE | pre_LcS | Yes | Yes | Yes | Yes | Yes |
| 9 | C | Yes | Radboudumc | MCE | pre_LcS | Yes | Yes | Yes | Yes | No |
| 10 | C | No | Radboudumc | MCE | pre_LcS | Yes | No | No | No | No |
| 11 | C | Yes | Radboudumc | MCE | pre_LcS | Yes | Yes | Yes | Yes | Yes |
| 12 | C | Yes | Radboudumc | MCE | pre_LcS | Yes | Yes | Yes | Yes | No |
| 13 | C | Yes | Radboudumc | MCE | pre_LcS | Yes | Yes | Yes | Yes | Yes |
| 14 | C | Yes | Radboudumc | MCE | pre_LcS | Yes | Yes | Yes | Yes | Yes |
| 15 | C | Yes | Radboudumc | MCE | pre_LcS | Yes | Yes | Yes | Yes | Yes |
| 16 | C | Yes | Radboudumc | MCE | pre_LcS | Yes | Yes | Yes | Yes | Yes |
| 17 | C | Yes | Radboudumc | MCE | pre_LcS | Yes | Yes | Yes | Yes | Yes |
| 18 | C | Yes | Radboudumc | MCE | pre_LcS | Yes | Yes | Yes | Yes | Yes |
| 19 | C | Yes | Radboudumc | MCE | pre_LcS | Yes | Yes | Yes | Yes | Yes |
| 21 | C | Yes | Bernhoven | MCE | pre_LcS | Yes | Yes | Yes | Yes | Yes |
| 22 | C | Yes | Bernhoven | MCE | pre_LcS | Yes | Yes | Yes | Yes | Yes |
| 23 | C | Yes | Bernhoven | MCE | pre_LcS | Yes | Yes | Yes | Yes | Yes |
| 24 | C | No | Bernhoven | MCE | pre_LcS | Yes | No | No | No | No |
| 25 | C | Yes | Bernhoven | MCE | pre_LcS | Yes | Yes | Yes | Yes | Yes |
| 26 | C | Yes | Radboudumc | MCE | pre_LcS | Yes | Yes | Yes | Yes | Yes |
| 1 | D | Yes | Radboudumc | MCE | post_LcS | No | No | No | Yes | No |
| 2 | D | Yes | Radboudumc | MCE | post_LcS | No | No | No | Yes | Yes |
| 3 | D | Yes | Radboudumc | MCE | post_LcS | No | No | No | Yes | Yes |
| 4 | D | Yes | Radboudumc | MCE | post_LcS | No | No | No | Yes | Yes |
| 5 | D | Yes | Radboudumc | MCE | post_LcS | No | Yes | Yes | Yes | Yes |
| 6 | D | Yes | Radboudumc | MCE | post_LcS | No | Yes | Yes | Yes | Yes |
| 7 | D | Yes | Radboudumc | MCE | post_LcS | No | Yes | Yes | Yes | Yes |
| 8 | D | Yes | Radboudumc | MCE | post_LcS | No | Yes | Yes | Yes | Yes |
| 9 | D | Yes | Radboudumc | MCE | post_LcS | No | Yes | Yes | Yes | No |
| 10 | D | No | Radboudumc | MCE | post_LcS | No | No | Yes | No | No |
| 11 | D | Yes | Radboudumc | MCE | post_LcS | No | Yes | Yes | Yes | Yes |
| 12 | D | Yes | Radboudumc | MCE | post_LcS | No | Yes | Yes | Yes | No |
| 13 | D | Yes | Radboudumc | MCE | post_LcS | No | Yes | Yes | Yes | Yes |
| 14 | D | Yes | Radboudumc | MCE | post_LcS | No | Yes | Yes | Yes | Yes |
| 15 | D | Yes | Radboudumc | MCE | post_LcS | No | No | Yes | Yes | Yes |
| 16 | D | Yes | Radboudumc | MCE | post_LcS | No | Yes | Yes | Yes | Yes |
| 17 | D | Yes | Radboudumc | MCE | post_LcS | No | No | No | Yes | Yes |
| 18 | D | Yes | Radboudumc | MCE | post_LcS | No | Yes | Yes | Yes | Yes |
| 19 | D | Yes | Radboudumc | MCE | post_LcS | No | No | No | Yes | Yes |
| 21 | D | Yes | Bernhoven | MCE | post_LcS | No | Yes | Yes | Yes | Yes |
| 22 | D | Yes | Bernhoven | MCE | post_LcS | No | Yes | Yes | Yes | Yes |
| 23 | D | Yes | Bernhoven | MCE | post_LcS | No | Yes | Yes | Yes | Yes |
| 24 | D | No | Bernhoven | MCE | post_LcS | No | No | Yes | No | No |
| 25 | D | Yes | Bernhoven | MCE | post_LcS | No | Yes | Yes | Yes | Yes |
| 26 | D | Yes | Radboudumc | MCE | post_LcS | No | Yes | Yes | Yes | Yes |

FFPE= Formalin Fixed Paraffin Embedded; MCPE= Methacarn Fixed Paraffin Embedded, GH= Guanidine Hydrochloride stabilized. Patient samples displayed in red font are excluded per exclusion criteria as described in Material and methods.

#### Supplemental Table S2. Fluorescence *in situ* hybridization probes

|  | Target | Oligo Name-Label | Target sequence (5’-3’) |
| --- | --- | --- | --- |
| Panel mixture | *Bacteroidetes* | CFB719-Atto488 | AGCTGCCTTCGCAATCGG |
|  | *Firmicutes* | LGC354A-Cy3  LGC354B-Cy3  LGC354C-Cy3 | TGGAAGATTCCCTACTGC  CGGAAGATTCCCTACTGC  CCGAAGATTCCCTACTGC |
|  | *Gammaproteobacteria* | Gam42a-Cy5 | GCCTTCCCACATCGTTT |
| All bacteria | Universal Eubacteria | EUB338-Cy5Cy5 | GCTGCCTCCCGTAGGAGT |
| Negative control | complimentary to universal Eubacteria | Non-EUB338-Cy5Cy5 | ACTCCTACGGGAGGCAGC |

#### Supplemental Table S3. Post-hoc pairwise comparisons of alpha diversity

| **Experimental groups** | **Statistics** | **p-values** | **FDR** |
| --- | --- | --- | --- |
| NSE vs MCE | 1468.0 | 4.00E-06 | **4.00E-06** |
| pre_LcS vs post_LcS | 297.0 | 5.32E-09 | **5.32E-09** |
| NSE_pre_LcS vs NSE_post_LcS | 40 | 1.49E-06 | **2.99E-06** |
| NSE_pre_LcS vs MCE_pre_LcS | 439 | 4.19E-07 | **1.26E-06** |
| NSE_pre_LcS vs MCE_post_LcS | 185 | 1.90E-01 | 1.90E-01 |
| NSE_post_LcS vs MCE_pre_LcS | 457 | 1.46E-11 | **8.74E-11** |
| NSE_post_LcS vs MCE_post_LcS | 387 | 6.14E-05 | **7.37E-05** |
| MCE_pre_LcS vs MCE_post_LcS | 69 | 4.82E-06 | **7.23E-06** |

Post-hoc pairwise comparisons of alpha-diversity for all group comparisons at the genus level. Significant (p < 0.05) results are in bold.

**Supplemental Table S4:** **Pairwise PERMANOVA**

| Experimental groups | F-value | R-square | p-values | FDR |
| --- | --- | --- | --- | --- |
| NSE vs MCE | 17.351 | 0.16953 | 0.001 | **0.001** |
| pre_LcS vs post_LcS | 13.133 | 0.13383 | 0.001 | **0.001** |
| MCE_pre_LcS vs MCE_post_LcS | 7.2402 | 0.1413 | 0.008 | **0.008** |
| NSE_pre_LcS vs MCE_pre_LcS | 7.1909 | 0.14618 | 0.002 | **0.0024** |
| NSE_pre_LcS vs NSE_post_LcS | 14.429 | 0.27007 | 0.001 | **0.0015** |
| NSE_pre_LcS vs MCE_post_LcS | 25.568 | 0.3784 | 0.001 | **0.0015** |
| NSE_post_LcS vs MCE_pre_LcS | 13.309 | 0.24506 | 0.001 | **0.0015** |
| NSE_post_LcS vs MCE_post_LcS | 17.454 | 0.2986 | 0.001 | **0.0015** |

Summary of multivariate pairwise PERMANOVA testing for statistical differences between esophageal regions and interventions. Significant (p < 0.05) results are in bold.

**Supplemental table S5:** Rank correlation analysis using Linear discriminant analysis effect size (LEfSe) at the genus level between the esophageal regions normal squamous (NSE) and metaplastic columnar epithelium (MCE)

| **Taxa (Phylum)** | **P-values^#^** | **FDR^§^** | **NSE*** | **MCE*** | **LDA score^** |
| --- | --- | --- | --- | --- | --- |
| Actinobacteria | 9.77E-03 | 2.20E-02 | 38135 | 62894 | 4.09 |
| Fusobacteria | 2.54E-02 | 3.81E-02 | 1333.7 | 6815.3 | 3.44 |
| Patescibacteria | 2.05E-04 | 6.15E-04 | 253.38 | 663.69 | 2.31 |
| Epsilonbacteraeota | 2.21E-02 | 3.81E-02 | 765.91 | 974.88 | 2.02 |
| Cyanobacteria | 1.49E-15 | 6.71E-15 | 366.38 | 54.789 | -2.2 |
| Acidobacteria | 3.53E-16 | 3.18E-15 | 1249.6 | 224.41 | -2.71 |
| **Taxa (Genus)** | **P-values** | **FDR** | **NSE** | **MCE** | **LDA score** |
| *Staphylococcus_sp__clone_MN2901_E2* | 1.07E-15 | 2.30E-14 | 5448.6 | 59874 | -4.43 |
| *Reyranella* | 3.00E-15 | 4.30E-14 | 11567 | 44842 | -4.22 |
| *Sphingomonas* | 1.86E-07 | 6.41E-07 | 27989 | 53864 | -4.11 |
| *Staphylococcus* | 1.07E-15 | 2.30E-14 | 695.67 | 14118 | -3.83 |
| *Bradyrhizobium_sp__SO_226* | 2.47E-05 | 6.85E-05 | 21942 | 31528 | -3.68 |
| *Chitinophaga* | 1.07E-15 | 2.30E-14 | 1689.8 | 11054 | -3.67 |
| *Pseudolabrys* | 2.13E-15 | 3.67E-14 | 2900.2 | 8318.9 | -3.43 |
| *Hyphomicrobium* | 9.84E-05 | 2.49E-04 | 4519.1 | 6573 | -3.01 |
| *Bradyrhizobium* | 7.14E-13 | 4.73E-12 | 873.85 | 1556.2 | -2.53 |
| *Cutibacterium* | 1.91E-12 | 1.18E-11 | 389.01 | 1036.2 | -2.51 |
| *Arthrobacter* | 1.75E-08 | 6.84E-08 | 0 | 465.65 | -2.37 |
| *Hydrogenophilus* | 2.59E-07 | 8.57E-07 | 44.427 | 354.66 | -2.19 |
| *Anoxybacillus* | 1.14E-13 | 9.78E-13 | 19.773 | 245.84 | -2.06 |
| *Comamonas* | 3.17E-09 | 1.36E-08 | 3.9851 | 217.15 | -2.03 |
| *Rhodoplanes* | 3.61E-08 | 1.35E-07 | 62.544 | 265.24 | -2.01 |
| *Caulobacter* | 5.53E-11 | 2.97E-10 | 97.773 | 273.7 | -1.95 |
| *Hephaestia* | 4.33E-14 | 4.13E-13 | 87.631 | 251.63 | -1.92 |
| *Corynebacterium_1* | 1.07E-03 | 2.36E-03 | 55.872 | 175.53 | -1.78 |
| *Faecalibacterium* | 5.97E-08 | 2.14E-07 | 0.28439 | 97.674 | -1.7 |
| *Bryobacter* | 1.24E-09 | 5.59E-09 | 15.144 | 107.91 | -1.68 |
| *Streptomyces* | 2.68E-06 | 8.24E-06 | 203.1 | 297.19 | -1.68 |
| *Rudaea* | 3.91E-04 | 9.10E-04 | 170.38 | 259.65 | -1.66 |
| *Edaphobacter* | 7.39E-16 | 2.3E-14 | 4.3469 | 87.947 | -1.63 |
| *uncultured_cyanobacterium* | 6.10E-10 | 2.91E-09 | 28.751 | 107.68 | -1.61 |
| *Pseudomonas* | 1.83E-04 | 4.50E-04 | 200.55 | 275.29 | -1.58 |
| *Legionella* | 1.92E-10 | 9.73E-10 | 36.03 | 103.2 | -1.54 |
| *Roseomonas* | 3.66E-05 | 9.54E-05 | 48.846 | 0 | 1.41 |
| *TM7_phylum_sp__oral_clone_DR034* | 6.50E-04 | 1.47E-03 | 578.74 | 252.82 | 2.21 |
| *Ralstonia* | 1.21E-08 | 4.96E-08 | 678.28 | 313.35 | 2.26 |
| *Pseudonocardia* | 1.35E-03 | 2.83E-03 | 1265.6 | 889.81 | 2.28 |
| *Pseudaminobacter* | 1.20E-03 | 2.57E-03 | 1551.3 | 1031.6 | 2.42 |
| *Acinetobacter* | 3.39E-13 | 2.65E-12 | 1149.2 | 29.247 | 2.75 |
| *Sphingobacterium* | 3.37E-06 | 9.99E-06 | 1809.2 | 1.0876 | 2.96 |
| *Granulicatella* | 3.46E-05 | 9.29E-05 | 5226.1 | 2343.4 | 3.16 |
| *Actinomyces* | 2.56E-04 | 6.11E-04 | 6062 | 2724.3 | 3.22 |
| *Klebsiella* | 2.56E-11 | 1.47E-10 | 4264.2 | 793.29 | 3.24 |
| *Rothia* | 6.27E-07 | 2.00E-06 | 49322 | 16689 | 4.21 |
| Rank correlation analysis using (LEfSe) at the genus level between Pre and Post-LcS intervention | | | | | |
| **Taxa (Phylum)** | **P-values** | **FDR** | **pre_LcS** | **post_LcS** | **LDA score** |
| Firmicutes | 2.49E-05 | 7.47E-05 | 220070 | 311060 | 4.66 |
| Actinobacteria | 8.37E-07 | 7.53E-06 | 30354 | 72255 | 4.32 |
| Bacteroidetes | 8.03E-05 | 1.81E-04 | 31658 | 56680 | 4.1 |
| Epsilonbacteraeota | 1.27E-02 | 1.91E-02 | 464.76 | 1289.4 | 2.62 |
| Patescibacteria | 4.70E-04 | 8.46E-04 | 288.38 | 654.89 | 2.27 |
| Fusobacteria | 3.72E-02 | 4.79E-02 | 6709.9 | 1789.1 | -3.39 |
| Proteobacteria | 5.60E-06 | 2.52E-05 | 709610 | 555320 | -4.89 |
| **Taxa (Genus)** | **P-values** | **FDR** | **pre_LcS** | **post_LcS** | **LDA score** |
| *Mesorhizobium* | 8.48E-07 | 4.34E-05 | 285530 | 449910 | -4.91 |
| *Afipia* | 1.01E-06 | 4.34E-05 | 2247.7 | 3680.2 | -2.86 |
| *Pseudaminobacter* | 1.74E-05 | 3.74E-04 | 976.08 | 1629.1 | -2.52 |
| *Abiotrophia* | 5.61E-04 | 2.84E-03 | 200.62 | 107.59 | 1.68 |
| *Stomatobaculum* | 2.88E-03 | 1.12E-02 | 567.6 | 376.48 | 1.98 |
| *TM7_phylum_sp__oral_clone_DR034* | 3.87E-04 | 2.08E-03 | 570.48 | 283.12 | 2.16 |
| *Candidatus_Ancillula* | 7.51E-04 | 3.40E-03 | 614.11 | 261.68 | 2.25 |
| *Lachnoanaerobaculum* | 4.44E-05 | 5.32E-04 | 724.76 | 347.59 | 2.28 |
| *Lactobacillus* | 7.60E-05 | 6.42E-04 | 614.49 | 224.54 | 2.29 |
| *Leptotrichia* | 9.02E-04 | 3.88E-03 | 1300 | 813.08 | 2.39 |
| *Bifidobacterium* | 9.74E-05 | 6.42E-04 | 1282.7 | 346.24 | 2.67 |
| *Megasphaera* | 6.04E-04 | 2.89E-03 | 2252.5 | 1092.2 | 2.76 |
| *Haemophilus* | 2.19E-05 | 3.76E-04 | 2313.8 | 1132.5 | 2.77 |
| *Atopobium* | 6.81E-06 | 1.95E-04 | 3728.9 | 1469.8 | 3.05 |
| *Prevotella_6* | 9.41E-05 | 6.42E-04 | 4026.7 | 1361.5 | 3.13 |
| *Granulicatella* | 3.18E-05 | 4.56E-04 | 5445.6 | 2325.5 | 3.19 |
| *Actinomyces* | 5.72E-05 | 5.47E-04 | 6254.7 | 2763.6 | 3.24 |
| *Gemella* | 1.68E-03 | 6.89E-03 | 17223 | 12315 | 3.39 |
| *Veillonella* | 1.05E-04 | 6.42E-04 | 23825 | 9912.1 | 3.84 |
| *Prevotella_7* | 4.95E-05 | 5.32E-04 | 38655 | 12967 | 4.11 |
| *Rothia* | 2.37E-04 | 1.36E-03 | 52546 | 15763 | 4.26 |
| *Streptococcus* | 9.41E-05 | 6.42E-04 | 224240 | 157580 | 4.52 |

\# Kruskal-Wallis rank-sum test

§ Adjusted for false discovery rate (FDR) using the Benjamini–Hochberg procedure

* Read counts

^ Linear Discriminant Analysis (LDA) scores >2.5 or <−2.5 and an FDR-corrected p < 0.05 were considered significant.

### Supplemental references

1.	Sharma P, Dent J, Armstrong D, Bergman JJ, Gossner L, Hoshihara Y, et al. The development and validation of an endoscopic grading system for Barrett's esophagus: the Prague C & M criteria. Gastroenterology. 2006;131(5):1392-9.

2.	Abela JE, Going JJ, Mackenzie JF, McKernan M, O'Mahoney S, Stuart RC. Systematic four-quadrant biopsy detects Barrett's dysplasia in more patients than nonsystematic biopsy. The American journal of gastroenterology. 2008;103(4):850-5.

3.	Bruggeling CE, Garza DR, Achouiti S, Mes W, Dutilh BE, Boleij A. Optimized bacterial DNA isolation method for microbiome analysis of human tissues. Microbiologyopen. 2021;10(3):e1191.

4.	Wouters Y, Dalloyaux D, Christenhusz A, Roelofs HMJ, Wertheim HF, Bleeker-Rovers CP, et al. Droplet digital polymerase chain reaction for rapid broad-spectrum detection of bloodstream infections. Microb Biotechnol. 2020;13(3):657-68.

5.	Klaschik S, Lehmann LE, Raadts A, Book M, Hoeft A, Stuber F. Real-time PCR for detection and differentiation of gram-positive and gram-negative bacteria. J Clin Microbiol. 2002;40(11):4304-7.

6.	Fujimoto J, Matsuki T, Sasamoto M, Tomii Y, Watanabe K. Identification and quantification of Lactobacillus casei strain Shirota in human feces with strain-specific primers derived from randomly amplified polymorphic DNA. Int J Food Microbiol. 2008;126(1-2):210-5.

7.	Magoc T, Salzberg SL. FLASH: fast length adjustment of short reads to improve genome assemblies. Bioinformatics. 2011;27(21):2957-63.

8.	Bokulich NA, Subramanian S, Faith JJ, Gevers D, Gordon JI, Knight R, et al. Quality-filtering vastly improves diversity estimates from Illumina amplicon sequencing. Nat Methods. 2013;10(1):57-U11.

9.	Caporaso JG, Kuczynski J, Stombaugh J, Bittinger K, Bushman FD, Costello EK, et al. QIIME allows analysis of high-throughput community sequencing data. Nat Methods. 2010;7(5):335-6.

10.	Edgar RC, Haas BJ, Clemente JC, Quince C, Knight R. UCHIME improves sensitivity and speed of chimera detection. Bioinformatics. 2011;27(16):2194-200.

11.	Haas BJ, Gevers D, Earl AM, Feldgarden M, Ward DV, Giannoukos G, et al. Chimeric 16S rRNA sequence formation and detection in Sanger and 454-pyrosequenced PCR amplicons. Genome Res. 2011;21(3):494-504.

12.	Edgar RC. UPARSE: highly accurate OTU sequences from microbial amplicon reads. Nat Methods. 2013;10(10):996-+.

13.	Schloss PD, Westcott SL, Ryabin T, Hall JR, Hartmann M, Hollister EB, et al. Introducing mothur: Open-Source, Platform-Independent, Community-Supported Software for Describing and Comparing Microbial Communities. Appl Environ Microb. 2009;75(23):7537-41.

14.	Quast C, Pruesse E, Yilmaz P, Gerken J, Schweer T, Yarza P, et al. The SILVA ribosomal RNA gene database project: improved data processing and web-based tools. Nucleic Acids Res. 2013;41(D1):D590-D6.

15.	Edgar RC. MUSCLE: multiple sequence alignment with high accuracy and high throughput. Nucleic Acids Res. 2004;32(5):1792-7.

16.	White JR, Nagarajan N, Pop M. Statistical Methods for Detecting Differentially Abundant Features in Clinical Metagenomic Samples. Plos Comput Biol. 2009;5(4).

17.	Greuter D, Loy A, Horn M, Rattei T. probeBase--an online resource for rRNA-targeted oligonucleotide probes and primers: new features 2016. Nucleic Acids Res. 2016;44(D1):D586-9.
